# Medication Clusters at Hospital Discharge and Risk of Adverse Drug Events at 30-days Post-Discharge: A Population-based Cohort Study of Older Adults

**DOI:** 10.1101/2022.08.28.22279291

**Authors:** Daniala L. Weir, Xiaomeng Ma, Lisa McCarthy, Terence Tang, Lauren Lapointe-Shaw, Walter P. Wodchis, Olavo Fernandes, Emily G. McDonald

**Affiliations:** Division of Pharmacoepidemiology and Clinical Pharmacology, Department of Pharmaceutical Sciences, Utrecht University; Institute of Health Policy, Management and Evaluation, University of Toronto; Health System Performance Network, Institute of Health Policy, Management & Evaluation, University of Toronto; Institute for Better Health, Trillium Health Partners; Department of Pharmacy, Trillium Health Partners; Department of Internal Medicine, Trillium Health Partners; Department of Medicine, University of Toronto; University Health Network; Research Institute of the McGill University Health Centre, Montreal, Quebec, Canada; Clinical Practice Assessment Unit, Department of Medicine, McGill University, Montreal, Quebec, Canada

## Abstract

**Background:** Certain combinations of medications can be harmful and may lead to serious drug-drug interactions. Identifying potentially problematic medication clusters could help guide prescribing decisions in hospital.

**Objectives:** To characterize medication prescribing patterns at hospital discharge and determine which medication clusters are associated with an increased risk of adverse drug events (ADEs) in the 30-days post hospital discharge.

**Methods:** All residents of the province of Ontario in Canada aged 66 years or older admitted to hospital between March 2016-February 2017 were included. Identification of medication prescribing clusters at hospital discharge was conducted using latent class analysis. Cluster identification was based on medications dispensed 30-days post-hospitalization. Multivariable logistic regression was used to assess the potential association between membership to a particular medication cluster and ADEs post-discharge, while also evaluating other patient characteristics.

**Results:** 188,354 patients were included in the study cohort. Median age (IQR) was 77 (71-84) and patients had a median (IQR) of 9 (6-13) medications dispensed in the year prior to admission. The study population consisted of 6 separate clusters of dispensing patterns post discharge: Cardiovascular (14%), respiratory (26%), complex care needs (12%), cardiovascular and metabolic (15%), infection (10%) and surgical (24%). Overall, 12,680 (7%) patients had an ADE in the 30-days following discharge. After considering other patient characteristics, those in the respiratory cluster had the highest risk of ADEs (aOR: 1.12, 95% CI: 1.08-1.17) compared to all the other clusters, while those in the neurocognitive & complex care needs cluster had the lowest risk (aOR:0.82, 95% CI: 0.77-0.87).

**Conclusion:** This study suggests that ADEs post hospital discharge are linked to identifiable clusters of medications, in addition to non-modifiable patient characteristics, such as age and certain comorbidities. This information may help clinicians and researchers better understand what patient populations and which types of interventions may benefit patients, to reduce their risk of experiencing an ADE.

**KEY POINTS:** This study suggests that ADEs post hospital discharge are linked to identifiable clusters of medications, in addition to non-modifiable patient characteristics, such as age and certain comorbidities. This information may help clinicians and researchers better understand what patient populations and which types of interventions may benefit patients, to reduce their risk of experiencing an ADE.

**PLAIN LANGUAGE SUMMARY:** Certain combinations of medications prescribed to patients when they are being discharged from hospital can increase the risk of adverse events after hospital discharge.

## INTRODUCTION

Adverse drug events (ADEs) are common following an acute hospitalization, especially among older adults using multiple medications.1 Post-discharge ADEs arise from multiple factors including inappropriate or low value prescribing, medication changes by the in-hospital care team, side effects related to new medications, and inadequate monitoring of high risk medications (e.g., anti-coagulants, antihyperglycemics, or combinations thereof etc.).^2–4^ Medication changes occur often for older adults during hospitalization.^5–7^ For example, a recent study by our team found that medical and surgical patients discharged from hospital were prescribed an average of 8 medications at discharge with 4 medication changes; 61% of changes were newly prescribed medications, 24% were discontinuations, and 15% were dose changes.8 A related study demonstrated that newly prescribed medications were associated with the highest risk of ADEs following discharge, compared to medications that were continued from prior to admission.3

Associations between specific medications or medication classes and adverse events following hospitalization have been evaluated (e.g., newly initiated antihypertensives are associated with an increased fall risk post-discharge);^1, 9,10^ however, no study has examined the association between *clusters* of medications prescribed at discharge from hospital and the risk of ADEs to our knowledge. A medication cluster refers to groups of medications that a patient is taking concurrently over a given period of time. For example, the co-occurence of psychiatric disorders with diabetes, hypertension, and coronary artery disease within a single patient is common,^1^^1^ however, the combined safety of medications to treat these conditions simultaneously (e.g., combination of antihyperglycemics, calcium channel blockers, lipid modifying agents and psychoanaleptics) is not known.^12, 13^ Indeed, certain combinations of medications can be harmful and may lead to serious drug-drug and drug-condition interactions (e.g., opioids and gabapentinoids; combination sedatives; anticholinergic burden; combination blood thinners).^14, 15^ Identifying potentially problematic clusters could help guide prescribing decisions in hospital related to re-evaluating newly started therapies as well as potentially deprescribing usual home medications. This could also help identify patients at high risk of ADEs who would benefit from targeted pharmacy specialty support prior to discharge.

Using administrative health data, the primary objective of this study was to characterize medication prescribing patterns at hospital discharge and determine which medication clusters were associated with an increased risk of ADEs in the 30-days post hospital discharge. The secondary objective was to confirm other previously described patient-level factors associated with the occurrence of ADEs.

## METHODS

### Study design & setting

We conducted a population based retrospective cohort study from March 1, 2016-February 28, 2017, of residents aged 66 years of age or older living in the province of Ontario, Canada. Ontario has a diverse, multicultural population of approximately 2 million people aged 65 years of age or older. All Ontario residents receive universal access to physician and hospital services, and those aged 65 years and up receive medication coverage through a provincial publicly-funded drug insurance program. Ethics approval for data access was obtained from the Ontario Ministry of Health and Trillium Health Partners. Patient consent was not required since the data was fully anonymized.

### Data sources

Multiple linked healthcare administrative databases were utilized for this study. Patient demographics and healthcare records were linked using encrypted health card numbers which served as unique and anonymous identifiers across databases. The Ontario Registered Persons Database contains basic demographic information including death.^16^ The Ontario Drug Benefits database contains prescription medication claims for all residents aged 65 years or older for medications dispensed to non-hospitalized patients.^17^ The Canadian Institutes of Health Information’s Discharge Abstract Database and National Ambulatory Care Reporting System contains detailed information for all admissions to hospital and emergency department visits. The Ontario Health Insurance Plan physician claims database contains information on all outpatient services provided by fee-for-service physicians.

### Study Population

All Ontario residents aged 66 years or older admitted to hospital between March 1 2016-February 28 2017 were included. If patients were admitted more than once during the study period, only their first hospital episode was considered. We limited the study population to those aged 66 and above as medications are covered under the Ontario Drug Benefits (ODB) program for those aged 65 and above in Ontario, and thus outpatient medication dispensing data is available for this population from 1 year prior to hospitalization. We included only those patients who were discharged to home or to a long term care facility. Patients who were transferred to or from another acute care setting were excluded since we would not have a record of their most recent medication use histories. Additionally, we excluded patients who did not have at least one medication with a valid Anatomical Therapeutic Chemical (ATC) code dispensed in the 30-days following discharge prior to another healthcare visit. To improve the performance of our clustering algorithm and identify the most clinically relevant clusters, medication classes with a prevalence of less than 1% were excluded (see Appendix Table A1 for list of excluded medications) (Figure 1). Study patients were followed until they were re-hospitalized, visited the emergency department (ED), died from any cause or the study period ended (30-days following index date), whichever occurred first.

### Medications dispensed post-hospitalization

Medications dispensed following hospital discharge were measured using prescription claims for medications in the 30-days following hospital discharge. For each dispensed medication, information about the drug identification number (DIN), Anatomical Therapeutic Chemical (ATC code), dispensing date, quantity dispensed and days supply was available. Medications dispensed following a post-discharge healthcare encounter (outpatient visit, ED visit, hospital readmission following index date) were excluded to ensure dispensed medications would capture prescribing decisions made during the index hospital stay alone. This approach was intended to capture medications that were part of the patients’ discharge prescription (either newly prescribed or continued from the community prior to the index admission). New medications were defined as those which were not dispensed in the year prior to the index admission and were subsequently dispensed in the 30-days following discharge, while medications continued at hospital discharge were those that were dispensed in the 1-year prior to admission and also dispensed in the 30-days following discharge. Medication class was defined using 3-digit ATC codes^18^ and distinct medications within each class were identified according to generic drug name.

### Patient characteristics potentially associated with ADEs

We measured a number of patient characteristics informed by previously published studies on risk factors for ADEs in the hospital and post-discharge setting^9, 19–21^ and included: patient demographics (age, sex), health service utilization in the one year prior to hospitalization (number of hospitalizations, ED visits, distinct medications dispensed), pre-admission chronic conditions (identified using ICD-9 & ICD-10 codes; appendix Table A2), number of pre-admission chronic conditions, characteristics of the hospital stay (institution admitted from, reason for admission, main patient service, discharge disposition), the number of medications dispensed overall in the 30-days following discharge as well as the number of new medications.

### Outcomes

The primary outcome for this study was the occurrence of ADEs (identified using ICD-10 codes for ED visits and hospital readmissions; Appendix Table A3)^22^ in 30-days following the index hospital discharge. Discharge date for the first eligible hospital episode was the index date.

## STATISTICAL ANALYSIS

Descriptive statistics were used to summarize patient characteristics for the entire study cohort. To characterize medication use in the 30-days following discharge, the prevalence of each medication class was calculated (overall and incidence of new use) and the most common drugs in each class were identified.

Identification of medication prescribing clusters at hospital discharge was conducted using latent class analysis (LCA). LCA is a model-based method of clustering that identifies unique groups or segments in a population when we do not have a variable that records group membership.^23^ Cluster identification was based on medications dispensed 30-days post-hospitalization using medication classes (3-digit ATC codes). Each medication class was included as a binary variable at the patient level (patient dispensed a medication in this class yes/no). LCA was performed successively where we started with a single class model under the assumption that all patients were dispensed the same combinations of medications following discharge. Then, increasing numbers of classes were added one at a time. The optimal number of subgroups (“n”) was decided by considering the best fit for the data (identified using the Bayesian information criterion [BIC]) and clinical interpretability of each cluster. The best fitting model with n-classes was identified and each patient was assigned to their most likely class based on these model probabilities. The difference in the prevalence of each medication class within a particular cluster vs the prevalence of medication class in the overall population was calculated (e.g., prevalence of patients dispensed diuretics in cluster 1-prevalence of patients dispensed diuretics overall) as was the prevalence of cluster membership among those dispensed each medication class (e.g., number of patients in cluster 1 dispensed diuretics/total number of patients dispensed diuretics). Clinicians on the team reviewed each cluster and agreed upon a clinically meaningful description for each. Patient descriptives were calculated according to medication cluster membership. The crude risk of ADEs were calculated overall and for each cluster separately.

Multivariable logistic regression models were constructed to assess the potential association between membership to a particular medication cluster and ADEs post-discharge, while also evaluating other patient characteristics historically associated with ADEs. The binary outcome was the occurrence of at least one ADE in the 30-days following discharge. Both cluster membership and patient characteristics were included as covariates in the same model. Six separate logistic models were constructed where a binary variable was included for each cluster separately in order to obtain the adjusted risk associated with belonging to a given cluster compared to all other clusters. Adjusted odds ratios and 95% confidence intervals were obtained for each covariate in the logistic regression model.

### Sensitivity analysis

While ICD-10 code sets to identify ADE’s have high specificity, they only have a sensitivity of up to 28%.^24–26^ All cause events will capture events that medications likely did not contribute to (e.g., pneumonia, sepsis, urinary tract infection), however, ADEs can also manifest as worsening of a patient’s underlying disease (e.g., statin non-adherence and occurrence of stroke)^27^ which would not be coded as being medication related. Therefore, in order to confirm the robustness of our results, and capture adverse events that were not necessarily coded as drug related, we re-conducted our primary analysis using a composite outcome of all cause emergency department visits or mortality in 30-days.

## RESULTS

Overall, 837,655 patients were admitted to hospital in Ontario between March 1 2016 and February 28 2017. After excluding those who were younger than 66 (n=544,064; 65%), those who were admitted from another acute care facility (n=10,720; 1%), and patients who did not have eligible medications dispensed 30-days post discharge prior to another healthcare encounter (n=84,539; 10%), 188,354 patients were included in the study cohort (Figure 1).

Median age (IQR) of the study participants was 77 (71-84) years and 53% were female. In the 1-year prior to the index hospital stay, 18% of patients had at least one hospitalization, patients had a median (IQR) of 2 (1-3) ED visits, 9 (6-13) medications dispensed, and 2 (1-3) chronic conditions. The most common chronic conditions included cancer (32% of patients), diabetes (26%) and osteoarthritis (25%). Most patients were admitted from the community and were not receiving home care (84%), 11% were admitted from long-term care and 5% were admitted from an outpatient clinic. The most common services patients were admitted to included general medicine (43%), orthopedic surgery (15%), general surgery (10%) and cardiology (8%). In the 30-days following discharge, a median (IQR) of 3 medications (2-6) were dispensed overall and 79% of patients had at least one new medication dispensed (Table 1). The top 5 most common medication classes (including new and continued medications) included: analgesics (40% prevalence, most common were hydromorphone and oxycodone), antithrombotics (30% prevalence, most common warfarin and clopidogrel), lipid modifying agents (25% prevalence, most common atorvastatin), drugs for acid related disorders (24% prevalence, most common pantoprazole) and antibacterials for systemic use (23% prevalence, most common ciprofloxacin, cefalexin, amoxicillin clavulanic acid cefuroxime) (Table 2).

**Table 1.**
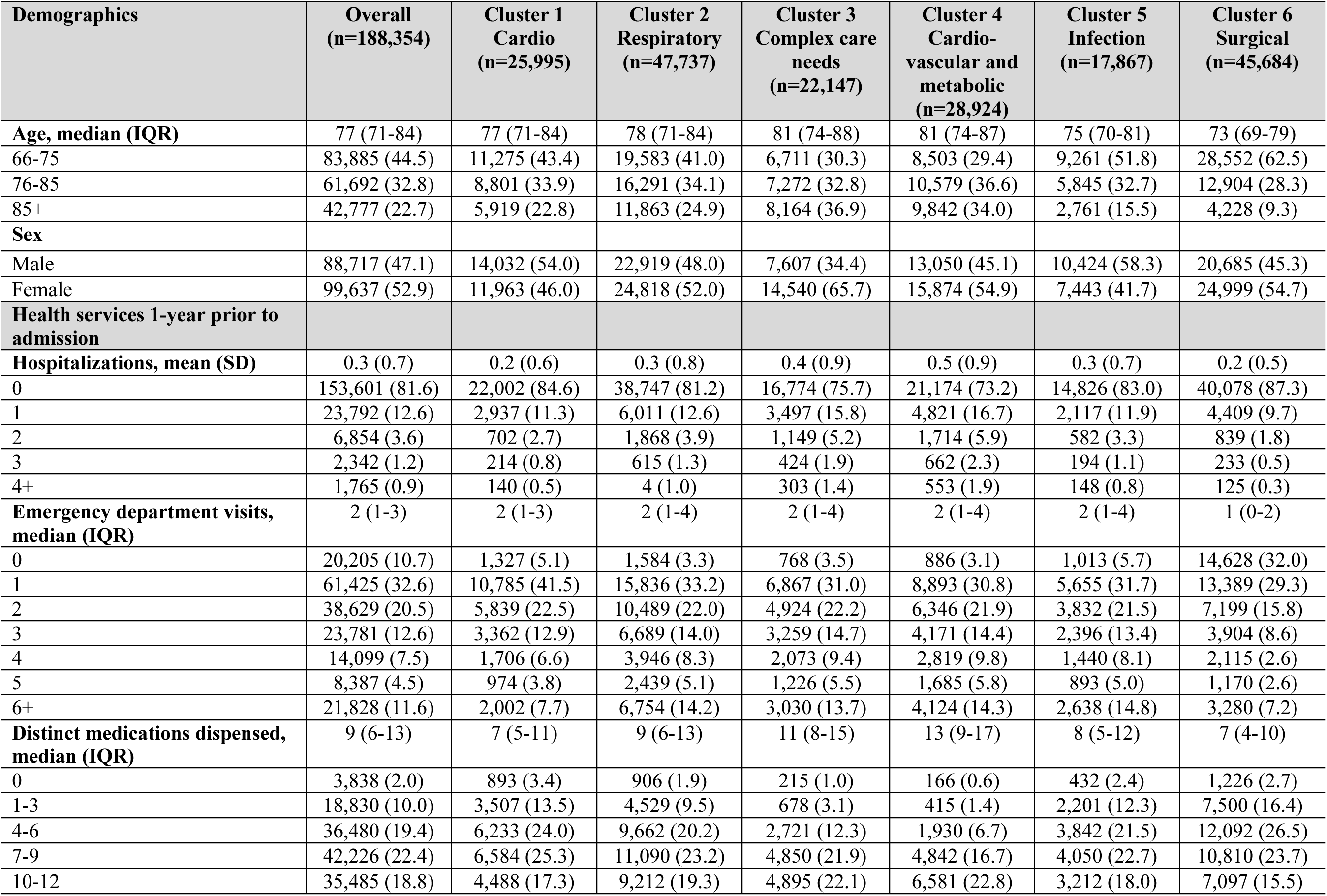
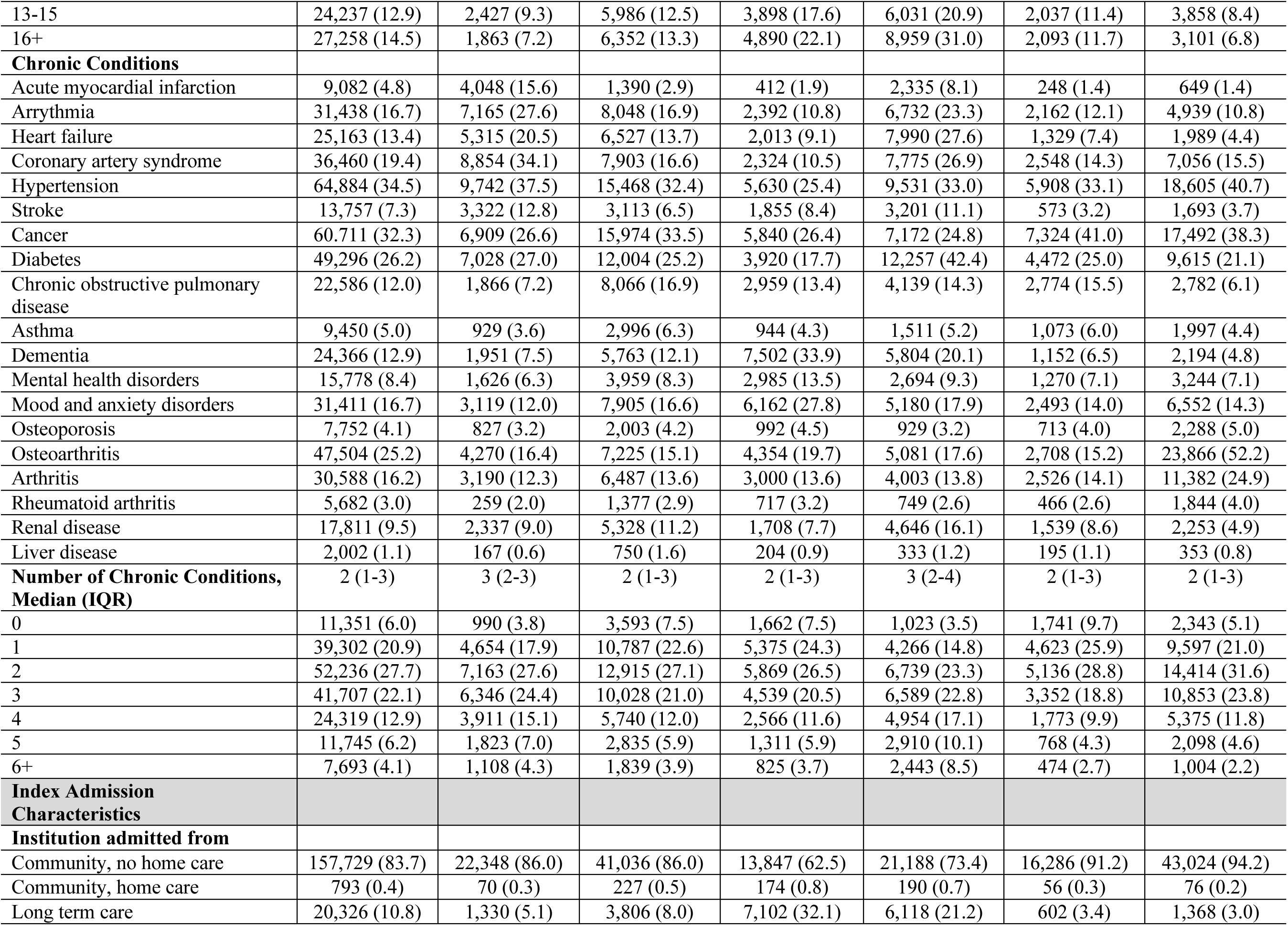
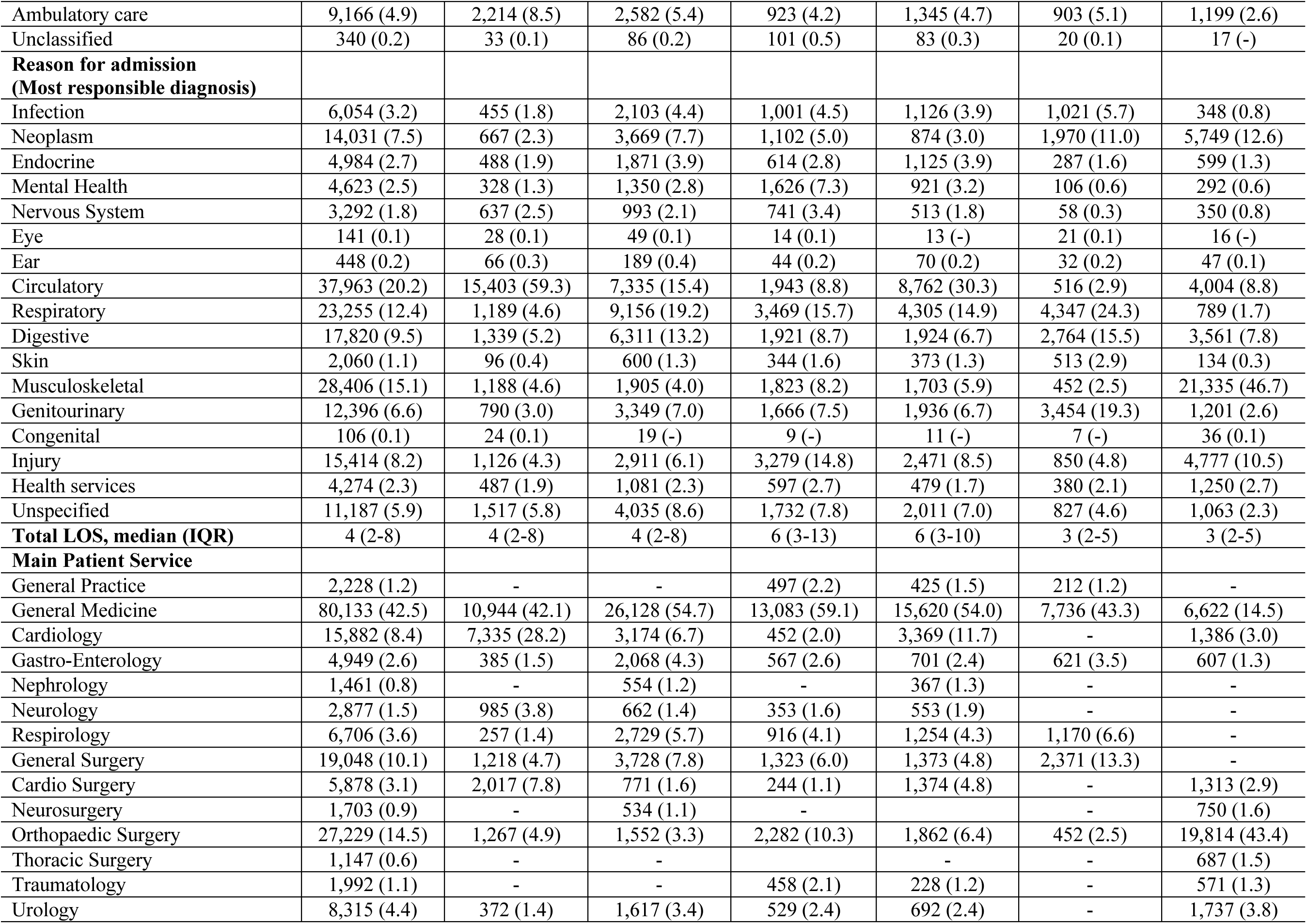
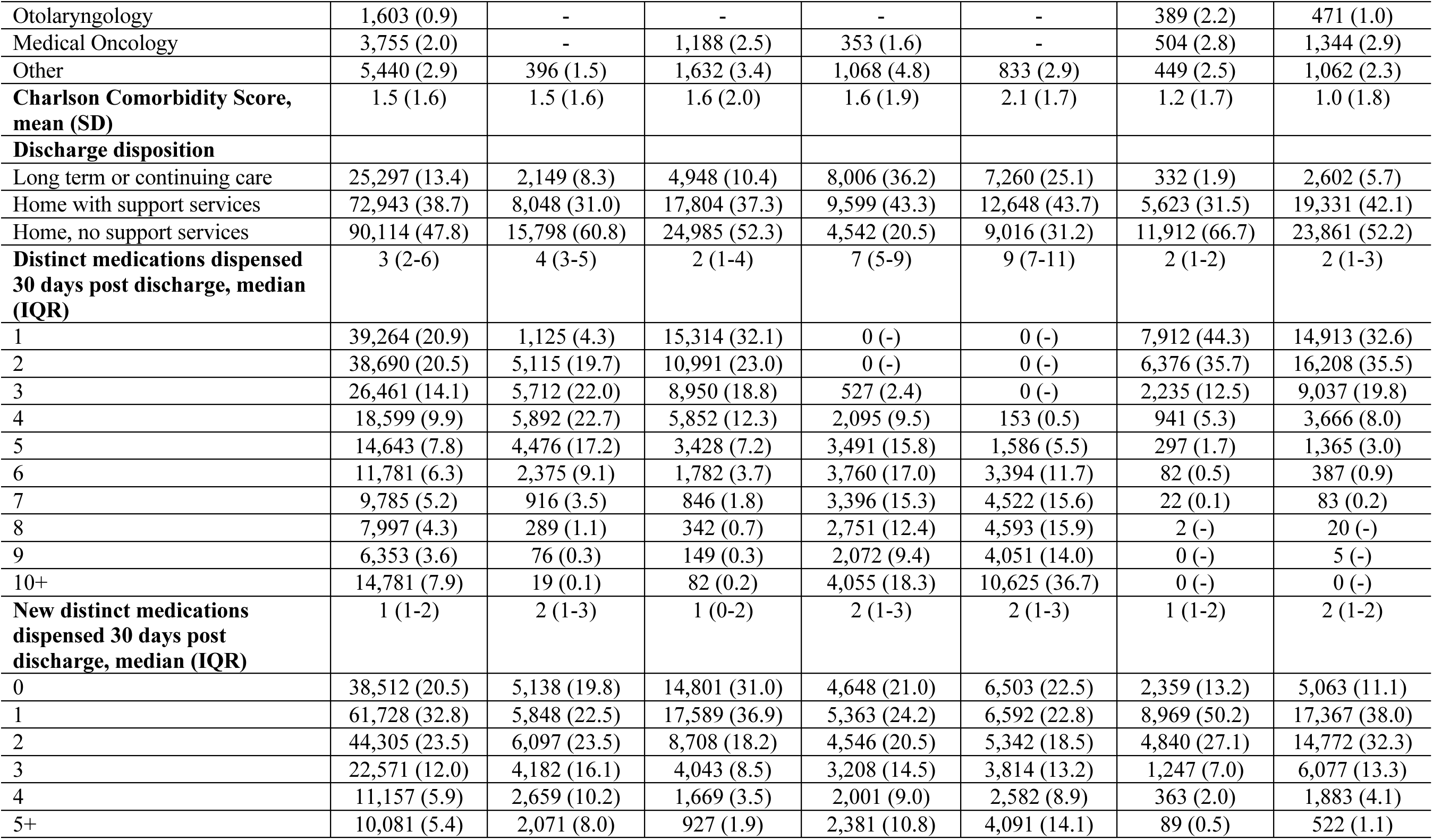
Patient characteristics overall and according to cluster

**Table 2.**
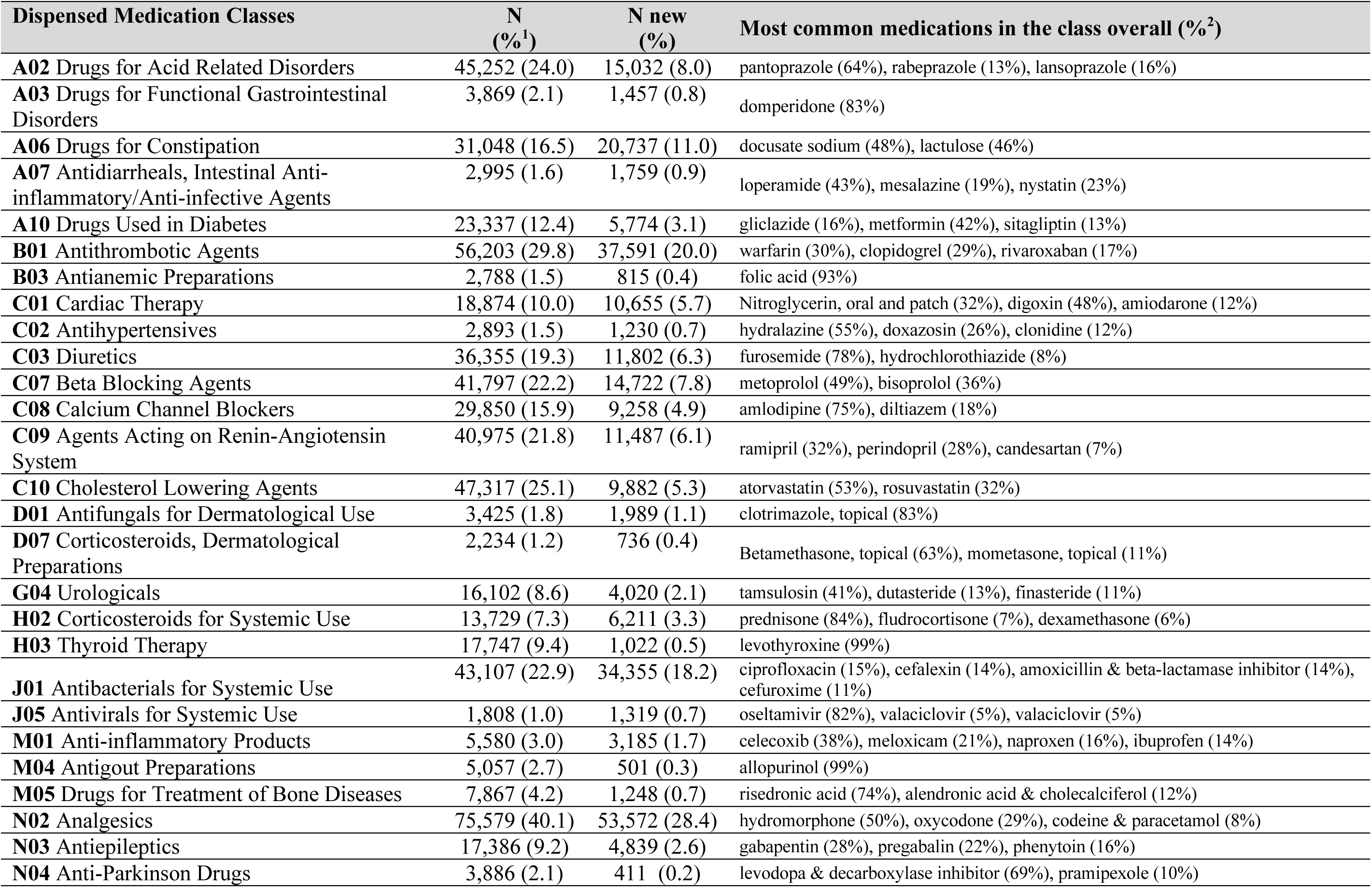
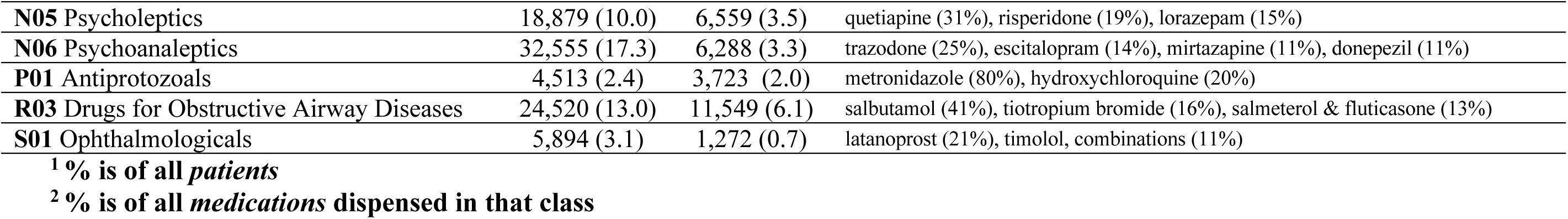
Prevalence of dispensed medication classes and most common ingredients in class

Goodness of fit statistics indicated that the study population consisted of 6 separate clusters of medication dispensing patterns. **Cluster 1** (“cardiovascular” cluster, n=25,995; 14% of study cohort) was characterized by use of beta blocking agents (e.g., metoprolol), cholesterol lowering agents (e.g., atorvastatin), antithrombotics (e.g., warfarin, clopidogrel), agents acting on the renin angiotensin system (e.g., ramipril, perindopril) and cardiac therapy (e.g., digoxin, nitroglycerin). Patients in this cluster were more likely to be male, had a higher prevalence of acute myocardial infarction, arrhythmia, heart failure and coronary artery syndrome, were more likely to be admitted from home without homecare, to be admitted for a circulatory related diagnosis, and receive care on a cardiology unit (Table 1). **Cluster 2** (“respiratory” cluster, n=47,747; 26%) was characterized by use of corticosteroids for systemic use (e.g.,prednisone) drugs for obstructive airway diseases (e.g, salbutamol) and antivirals for systemic use (e.g., oseltamivir). Patients in this cluster had a higher prevalence of COPD and were more likely to have been admitted for a respiratory related diagnosis and to receive care on a general medicine unit (Table 1). **Cluster 3** (“complex care needs” cluster, n=22,147; 12%) was characterized by use of antidepressants (e.g., trazodone, escitalopram) antipsychotics (e.g., quetiapine, risperidone), drugs for acid related disorders (e.g., pantoprazole), gabapentinoids (e.g., gabapentin, pregabalin), analgesics (e.g., hydromorphone, oxycodone), drugs for constipation (e.g., docusate), thyroid therapy (e.g., levothyroxine) and anti-parkinson’s drugs (e.g., carbidopa/levodopa). Patients in this cluster were more likely to be female, had a greater number of medications dispensed prior to admission, were more likely to have dementia, any mental health disorder, or mood/anxiety disorder, were more likely to have been admitted from long term care, be admitted for a mental health, nervous system or injury related diagnosis and were most likely to receive care on a general medicine unit. (Table 1) **Cluster 4** (“cardiovascular and metabolic”; n=28,924, 15%) was characterized by use of cholesterol lowering agents, beta blocking agents, drugs for acid related disorders, diuretics (e.g., furosemide), agents acting on the renin-angiotensin system, drugs used in diabetes (e.g., metformin), calcium channel blockers (e.g., amlodipine), antipsychotics, anti-thrombotics, anti-gout preparations (e.g., allopurinol) and other anti-hypertensives (e.g., hydralazine). Patients in this cluster had more medications dispensed prior to admission, were more likely to have cardiovascular conditions, diabetes and renal disease, be admitted from long term care, be admitted for a circulatory related diagnosis, be on a general medicine or cardiology unit and had a high proportion of multiple new medications dispensed. (Table 1) **Cluster 5** (“infection” cluster; n=17,867, 10%) was characterized by use of antibacterials for systemic use (e.g., ciprofloxacin, cefalexin, amoxicillin & beta-lactamase inhibitors) and anti-fungals (e.g., metronidazole). Patients in this cluster were more likely to be male, have cancer, be admitted from home without homecare and be admitted for a malignancy, respiratory or digestive related diagnosis (Table 1). **Cluster 6** (“surgical” cluster; n=45,684, 24%) was characterized by use of analgesics, anti-thrombotics, drugs for constipation and anti-inflammatory products (e.g., celecoxib, meloxicam). Patients in this cluster were slightly younger, more likely to have osteoarthritis and arthritis, be admitted from home, be admitted for a malignancy, or musculoskeletal related diagnosis, and be on an orthopedic surgery unit (Table 1).

Overall, 12,680 (6.7%) patients had an ADE in the 30-days following discharge. The most common diagnosis associated with ADEs across the majority of clusters was acute kidney failure and volume depletion. Of note, enterocolitis due to *Clostridium difficile* was a common ADE in cluster 5 (infection) and pulmonary embolism in the absence of acute cardiorespiratory disease was common in cluster 6 (surgical). Crude risk of ADEs was highest for those in cluster 4 (cardiovascular and metabolic; 8.9%) and lowest for those in cluster 6 (surgical; 4.6%) (Table 3).

**Table 3.** Association Between Patient Characteristics and Risk of Adverse Drug Events in 30-days Following Discharge

| Patient Characteristics | Crude Risk of ADE (%) | Adjusted OR ADE (%) | Lower 95% CI for adjusted OR | Upper 95% CI for adjusted OR |
| --- | --- | --- | --- | --- |
| **Demographics** |  |  |  |  |
| **Age** |  |  |  |  |
| 66-75 | 5.7 |  | *Reference* |  |
| 76-85 | 7.3 | 1.13 | 1.09 | 1.19 |
| 85+ | 8.0 | 1.19 | 1.13 | 1.26 |
| **Sex** |  |  |  |  |
| Male | 6.7 |  | *Reference* |  |
| Female | 6.8 | 1.02 | 0.99 | 1.06 |
| **Health services 1-year prior to admission** |  |  |  |  |
| **Hospitalizations** |  |  |  |  |
| 0 | 6.2 |  | *Reference* |  |
| 1 | 7.9 | 1.00 | 0.94 | 1.06 |
| 2 | 10.3 | 1.08 | 0.99 | 1.18 |
| 3 | 12.7 | 1.22 | 1.07 | 1.39 |
| 4+ | 14.1 | 1.24 | 1.07 | 1.44 |
| **Emergency department visits** |  |  |  |  |
| 0 | 3.0 |  | *Reference* |  |
| 1 | 5.7 | 1.19 | 1.07 | 1.32 |
| 2 | 6.5 | 1.29 | 1.16 | 1.44 |
| 3 | 7.9 | 1.49 | 1.33 | 1.67 |
| 4 | 8.1 | 1.45 | 1.28 | 1.63 |
| 5 | 8.4 | 1.44 | 1.26 | 1.64 |
| 6+ | 10.6 | 1.69 | 1.50 | 1.90 |
| **Distinct medications dispensed** |  |  |  |  |
| 0 | 4.7 |  | *Reference* |  |
| 1-3 | 4.6 | 1.02 | 0.87 | 1.21 |
| 4-6 | 5.1 | 1.08 | 0.92 | 1.26 |
| 7-9 | 6.0 | 1.15 | 0.98 | 1.35 |
| 10-12 | 7.0 | 1.26 | 1.07 | 1.48 |
| 13-15 | 8.1 | 1.37 | 1.16 | 1.61 |
| 16+ | 10.2 | 1.55 | 1.31 | 1.83 |
| **Chronic Conditions** |  |  |  |  |
| Acute myocardial infarction | 8.7 | 1.03 | 0.95 | 1.12 |
| Arrhythmia | 8.6 | 1.10 | 1.05 | 1.16 |
| Heart failure | 11.6 | 1.21 | 1.15 | 1.27 |
| Coronary artery syndrome | 7.4 | 0.96 | 0.91 | 1.01 |
| Hypertension | 6.7 | 1.03 | 0.99 | 1.07 |
| Stroke | 7.2 | 0.92 | 0.94 | 0.99 |
| Cancer | 6.9 | 0.96 | 0.92 | 1.00 |
| Diabetes | 7.6 | 0.97 | 0.92 | 1.01 |
| Chronic obstructive pulmonary disease | 7.9 | 0.99 | 0.94 | 1.05 |
| Asthma | 7.5 | 1.01 | 0.93 | 1.09 |
| Dementia | 7.8 | 1.01 | 0.95 | 1.07 |
| Mental health disorders | 7.2 | 0.99 | 0.92 | 1.05 |
| Mood and anxiety disorders | 7.4 | 1.02 | 0.98 | 1.08 |
| Osteoporosis | 6.4 | 0.98 | 0.89 | 1.08 |
| Osteoarthritis | 5.3 | 0.99 | 0.94 | 1.05 |
| Arthritis | 6.2 | 0.97 | 0.92 | 1.02 |
| Rheumatoid arthritis | 6.7 | 1.00 | 0.90 | 1.12 |
| Renal disease | 11.6 | 1.42 | 1.34 | 1.50 |
| Liver disease | 12.6 | 1.54 | 1.34 | 1.77 |
| **Index admission characteristics** |  |  |  |  |
| **Institution admitted from** |  |  |  |  |
| Community, no home care | 6.5 |  | *Reference* |  |
| Community, home care | 7.9 | 0.91 | 0.70 | 1.18 |
| Long term care | 8.3 | 1.09 | 1.03 | 1.17 |
| Ambulatory care | 7.7 | 1.09 | 1.01 | 1.19 |
| Unclassified | 7.7 | 1.13 | 0.76 | 1.70 |
| **Acuity** |  |  |  |  |
| Not admitted from ED | 4.2 |  | *Reference* |  |
| Admitted from the ED | 7.7 | 1.23 | 1.14 | 1.33 |
| **Main Patient Service** |  |  |  |  |
| General Medicine | 8.1 |  | *Reference* |  |
| General Practice | 6.5 | 0.81 | 0.68 | 0.97 |
| Cardiology | 8.2 | 0.95 | 0.88 | 1.02 |
| Gastro-Enterology | 8.6 | 1.02 | 0.91 | 1.15 |
| Nephrology | 10.9 | 0.93 | 0.78 | 1.11 |
| Neurology | 6.5 | 0.91 | 0.77 | 1.07 |
| Respirology | 7.7 | 1.02 | 0.92 | 1.13 |
| General Surgery | 4.2 | 0.68 | 0.62 | 0.74 |
| Cardio Surgery | 6.9 | 1.06 | 0.94 | 1.20 |
| Neurosurgery | 5.2 | 0.98 | 0.78 | 1.23 |
| Orthopaedic Surgery | 3.6 | 0.84 | 0.75 | 0.94 |
| Thoracic Surgery | 6.0 | 0.89 | 0.69 | 1.15 |
| Urology | 4.2 | 0.60 | 0.53 | 0.68 |
| Otolaryngology | 3.9 | 0.64 | 0.49 | 0.83 |
| Medical Oncology | 8.7 | 0.95 | 0.83 | 1.09 |
| Other | 6.7 | 0.99 | 0.88 | 1.11 |
| **Reason for Admission\*** |  |  |  |  |
| Circulatory | 8.4 |  | *Reference* |  |
| Infection | 6.8 | 1.00 | 0.90 | 1.11 |
| Neoplasm | 8.4 | 1.08 | 0.97 | 1.20 |
| Endocrine | 7.5 | 0.95 | 0.85 | 1.07 |
| Mental Health | 7.5 | 0.87 | 0.77 | 0.99 |
| Nervous System | 5.8 | 0.78 | 0.67 | 0.91 |
| Eye | 4.3 | 0.65 | 0.29 | 1.48 |
| Ear | 5.4 | 0.81 | 0.53 | 1.24 |
| Respiratory | 7.2 | 0.76 | 0.70 | 0.83 |
| Digestive | 6.9 | 0.98 | 0.90 | 1.07 |
| Skin | 8.3 | 0.91 | 0.77 | 1.08 |
| Musculoskeletal | 3.8 | 0.86 | 0.77 | 0.96 |
| Genitourinary | 7.5 | 1.04 | 0.95 | 1.15 |
| Congenital | 3.8 | 0.68 | 0.25 | 1.84 |
| Injury | 4.5 | 0.64 | 0.58 | 0.71 |
| Health services | 4.9 | 0.65 | 0.56 | 0.75 |
| Unspecified | 7.2 | 0.93 | 0.86 | 1.01 |
| **Total length of stay** | - | 1.01 | 1.01 | 1.02 |
| **Charlson Comorbidity Score** | - | 1.05 | 1.04 | 1.06 |
| **Post-discharge medications** |  |  |  |  |
| **New Medications** |  |  |  |  |
| 0 | 6.7 | **Reference** |  |  |
| 1 | 6.7 | 1.14 | 1.08 | 1.21 |
| 2 | 6.2 | 1.16 | 1.09 | 1.22 |
| 3 | 6.8 | 1.23 | 1.15 | 1.32 |
| 4 | 7.5 | 1.31 | 1.20 | 1.42 |
| 5+ | 8.4 | 1.39 | 1.27 | 1.52 |
| **Medication Cluster <sup>1</sup>** |  |  |  |  |
| Cardiovascular | 7.0 | 0.97 | 0.92 | 1.03 |
| Respiratory | 7.7 | 1.12 | 1.08 | 1.17 |
| Complex care needs | 6.6 | 0.82 | 0.77 | 0.87 |
| Cardiovascular and metabolic | 8.9 | 0.98 | 0.94 | 1.03 |
| Infection | 6.0 | 1.01 | 0.94 | 1.08 |
| Surgical | 4.6 | 1.02 | 0.97 | 1.08 |

<sup>1</sup> Reference is all other clusters (e.g., cardiovascular vs not)

Our multivariable model included both variables indicating cluster membership as well as patient level variables, therefore, the interpretation of the odds ratio for a given covariate in the model is the risk of ADE’s associated with presence of a characteristic (vs reference) after accounting for the other patient characteristics included in the model. After considering other patient characteristics, those in the respiratory cluster had the highest risk of ADEs (adjusted odds ratio [aOR]: 1.12, 95% CI: 1.08-1.17) compared to all the other clusters while those in the neurocognitive & complex care needs cluster had the lowest risk (aOR:0.82, 95% CI: 0.77-0.87). There was no difference in the risk of ADEs among patients in the cardiovascular, metabolic syndrome, infection and surgical clusters when comparing each cluster to all other clusters. (Table 3)

Patient level factors that were also associated with an increased risk of ADEs included age, increasing healthcare service use in the year prior to admission (hospitalizations and ED visits), a greater number of pre-admission medications dispensed, presence of heart failure, arrhythmia, renal disease and liver disease, being admitted from long term care, being admitted from the ED, increasing length of stay, a higher Charlson Comorbidity Score, and a greater number of new medications dispensed in the 30-days following discharge (Table 3). Of all the variables in the model, belonging to the respiratory cluster, presence of renal disease, liver disease and heart failure were associated with the highest risk of ADEs.

Our sensitivity analyses indicated that the direction of the point estimates for the association between the characteristics included in our multivariable model and ED visits/mortality was similar when compared to the primary analysis. However, the magnitude of these associations was higher for the infection and surgical clusters. (Appendix Table A5).

## DISCUSSION

In this population-based retrospective cohort study of older hospitalized adults, we identified 6 clusters of medication prescribing patterns at hospital discharge; cardiovascular, respiratory, complex care needs, metabolic syndrome, infection and surgical clusters. After accounting for other factors known to increase the risk of ADE after discharge, patients in the respiratory cluster had the highest risk of an ADE after discharge while those in the complex care needs cluster had the lowest risk. We also confirmed that a number of previously reported patient characteristics were associated with an increased risk of ADEs, for example, the overall number of medications, the addition of new medications, as well as the presence of heart failure, renal disease and liver disease.

A systematic review published in 2018 aimed to determine risk factors for medication related harm among older adults after hospital discharge.1 Only 3 of the 8 included studies conducted multivariable analyses to evaluate risk factors for medication-related harm. Among these studies, only select medication classes (as opposed to combinations or clusters of medication classes) and diagnoses were included in the multivariable analyses. The majority of studies were of moderate quality and substantial methodological heterogeneity was identified with respect to how related harm was measured. A higher absolute number of medications,^28, 29^ the addition of new medications at discharge,^30^ warfarin^28, 29^, furosemide^31^, a lower Mini-Mental State Examination score^30^ and female sex^30^ were all independent risk factors for medication harm following hospitalization. Since this systematic review was published, three other studies have evaluated risk factors for medication-related harm following a hospital stay.^2, 9, 32^ Risk factors from these studies included the number of medication changes, prior hospitalizations preceding the index admission, older age, past adverse drug reaction(s), prescriptions for antiplatelet drugs, antidiabetic drugs, corticosteroids and again, a higher absolute number of medications were risk factors. Consistent risk factors for an increased risk of ADEs post hospital discharge are older age, the absolute number of medications and the addition of new medications at discharge, which we also confirmed in our analysis. However, we found no other study that specifically evaluated medication clusters or combinations of medications as an independent risk factor for ADEs post discharge.

The results of this study inform clinical practice in two important ways. First, this study provides evidence that medications tend to be prescribed in readily identifiable, predictable medication clusters. Furthermore, some clusters differentially impact the risk of post hospital ADEs independent of patient characteristics and the number of prescribed medications. Therefore, these results could inform clinicians about new potentially modifiable risk factors for ADEs, such as newly added combinations of medications, which may be important to review prior to the patient being discharged. For example, in the respiratory cluster, patients were most likely to be using a combination of corticosteroids, drugs for obstructive airway diseases and antivirals, and patients in this cluster also had the highest independent risk of ADEs post-discharge. Thus, medications prescribed to patients in this cluster (especially new medications) may be important to review prior to discharge, or patients may require counseling with respect to expected or predictable side effects from this medication cluster (e.g., gastrointestinal symptoms from systemic corticosteroids and oseltamivir, or delirium from combinations of anticholinergic inhalers and corticosteroids).

Second, similar to previous studies, better understanding of non-modifiable patient-level factors associated with an increased risk of ADEs in the 30-days post discharge (e.g., older age, presence of heart failure etc.) can inform clinicians which patients may benefit most from resource-intensive interventions to decrease ADEs (e.g., home care post discharge or targeted education related to signs and symptoms to monitor for related to a potential ADE.) For example, presence of heart failure, renal disease or liver disease could be important markers for patients at high risk of experiencing an ADE and should have their medications reviewed by a pharmacist prior to discharge. Interestingly, although the neurocognitive & complex care needs cluster was associated with the highest crude risk of ADEs, after considering other patient characteristics, this cluster was not associated with an increased risk of ADEs compared to any of the other clusters. This could indicate that these patients may be well known to be “high risk” and are receiving adequate support with respect to their medications in the post discharge period. A limitation of our study is that we did not have information on factors such as involvement of a pharmacist or consultation with geriatrics prior to discharge.

There are other limitations associated with this study that are important to keep in mind. First, although patients were dispensed particular medications, they might not have taken these medications as prescribed. This is a limitation associated with all studies which use dispensing data to capture drug exposure. Second, we did not have access to clinical data to include in our models (e.g., renal function, sodium levels) which may have led to unmeasured confounding. Third, the data analyzed in this study was from 2016/2017 and the prescribing patterns for certain medications have changed over this time period. For example, the use of direct oral anticoagulants has increased dramatically in the past 5 years. Lastly, although we analyzed both ADEs and emergency department visits as outcomes and found consistent results between the two models, it’s still important to acknowledge that use of ICD10 codes to measure ADEs will miss many outcomes which are drug related and ED visits will also capture a number of events which are not related to medications. Additional research is required with respect to valid measures of ADEs using administrative health data.

Despite these limitations, this study is an important step towards personalized medicine as it relates to appropriate prescribing. Identifying patients who are taking high risk clusters of medications could lead to interventions that improve the safety of medication use following hospitalization. By considering a patient’s whole drug regimen, in combination with their characteristics, we can take steps towards a more holistic approach to prescribing, not just focusing on treating single conditions or addressing single high risk drug classes in isolation.

In conclusion, this study suggests that ADEs post hospital discharge are linked to identifiable clusters of medications, in addition to non-modifiable patient characteristics, such as age and certain comorbidities. This information may help clinicians and researchers better understand what patient populations and which types of interventions may benefit patients, to reduce their risk of experiencing an ADE.

## Data Availability

The data referred to in the manuscript is not available upon request

## STATEMENTS AND DECLARATIONS

**Conflict of Interest:** Dr. McDonald receives salary support for research from the Fond de recherche santé Québec and is the owner and chief scientific officer of MedSafer, a software that helps guide clinicians through the process of deprescribing for older adults with polypharmacy (the intellectual property is jointly held with McGill University). Authors have no other competing interests to disclose.

## Author Contributions

Conceptualization - DW; Methodology - DW; Validation - DW, EGM; Formal Analysis - DW, XM; Investigation - All authors; Resources - DW, WPW; Data Curation - WPW; Writing - Original Draft - DW, EGM, LLS, TT, LM; Writing - Review and Editing - All authors; Visualization DW, EGM

**Sponsors Role:** N/A

---

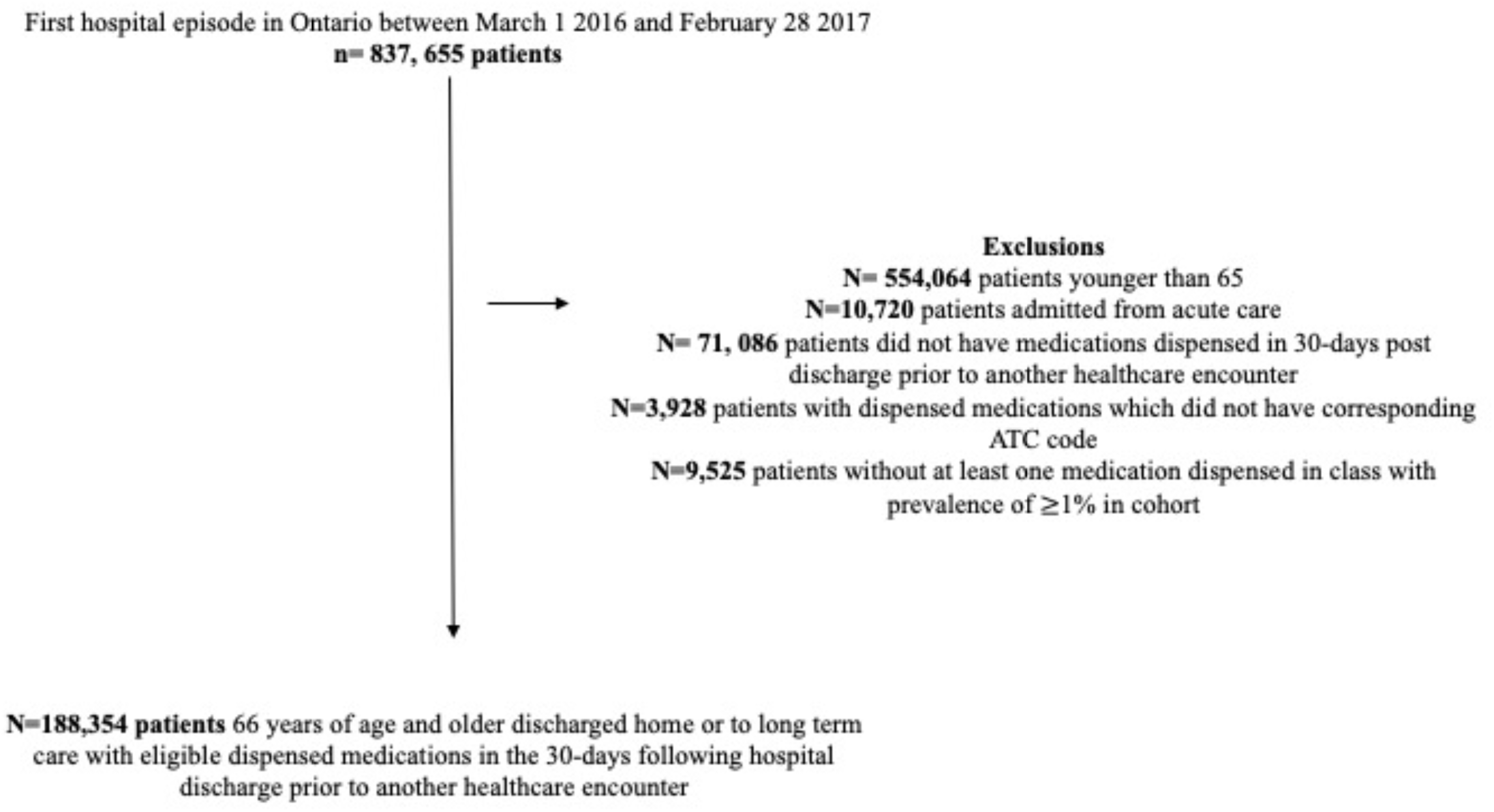

**Table A1.** List of medication classes excluded from analysis

| **3 Digit ATC code** | **Class Name** |
| --- | --- |
| A01 | STOMATOLOGICAL PREPARATIONS |
| A05 | BILE AND LIVER THERAPY |
| A09 | DIGESTIVES, INCL. ENZYMES |
| A11 | VITAMINS |
| A16 | OTHER ALIMENTARY TRACT AND METABOLISM PRODUCTS |
| B02 | ANTIHEMORRHAGICS |
| B05 | BLOOD SUBSTITUTES AND PERFUSION SOLUTIONS |
| C04 | PERIPHERAL VASODILATORS |
| C05 | VASOPROTECTIVES |
| D03 | PREPARATIONS FOR TREATMENT OF WOUNDS AND ULCERS |
| D05 | ANTIPSORIATICS |
| D06 | ANTIBIOTICS AND CHEMOTHERAPEUTICS FOR DERMATOLOGICAL USE |
| D10 | ANTI-ACNE PREPARATIONS |
| D11 | OTHER DERMATOLOGICAL PREPARATIONS |
| G01 | GYNECOLOGICAL ANTIINFECTIVES AND ANTISEPTICS |
| G02 | OTHER GYNECOLOGICALS |
| G03 | SEX HORMONES AND MODULATORS OF THE GENITAL SYSTEM |
| H01 | PITUITARY AND HYPOTHALAMIC HORMONES AND ANALOGUES |
| H04 | PANCREATIC HORMONES |
| H05 | CALCIUM HOMEOSTASIS |
| J02 | ANTIMYCOTICS FOR SYSTEMIC USE |
| J04 | ANTIMYCOBACTERIALS |
| J07 | VACCINES |
| L01 | ANTINEOPLASTIC AGENTS |
| L02 | ENDOCRINE THERAPY |
| L03 | IMMUNOSTIMULANTS |
| L04 | IMMUNOSUPPRESSANTS |
| M03 | MUSCLE RELAXANTS |
| N07 | OTHER NERVOUS SYSTEM DRUGS |
| P02 | ANTHELMINTICS |
| P03 | ECTOPARASITICIDES, INCL. SCABICIDES, INSECTICIDES AND REPELLENTS |
| R01 | NASAL PREPARATIONS |
| R05 | COUGH AND COLD PREPARATIONS |
| R06 | ANTIHISTAMINES FOR SYSTEMIC USE |
| R07 | OTHER RESPIRATORY SYSTEM PRODUCTS |
| S02 | OTOLOGICALS |
| S03 | OPHTHALMOLOGICAL AND OTOLOGICAL PREPARATIONS |
| V03 | ALL OTHER THERAPEUTIC PRODUCTS |
| V07 | ALL OTHER NON-THERAPEUTIC PRODUCTS |

**Table A2.** IC9 and ICD 10 codes used to define pre-admission chronic conditions

| Condition | ICD 9 | ICD 10 |
| --- | --- | --- |
| Acute Myocardial Infarction (AMI) | 410 | I21, I22 |
| **Osteo- and other Arthritis:** |  |  |
| (A) Osteoarthritis | 715 | M15-M19 |
| (B) Other Arthritis (includes Synovitis, Fibrositis, Connective tissue disorders, Ankylosing spondylitis, Gout Traumatic arthritis, pyogenic arthritis, Joint derangement, Dupuytren's contracture, Other MSK disorders) | 727, 729, 710, 720, 274, 716, 711, 718, 728, 739 | M00-M03, M07, M10, M11-M14, M20-M25, M30-M36, M65-M79 |
| Arthritis - Rheumatoid arthritis | 714 | M05-M06 |
| Asthma | 493 | J45 |
| (all) Cancers | 140-239 | C00-C26, C30-C44, C45-C97 |
| Cardiac Arrhythmia | 427 (OHIP) / 427.3 (DAD) | I48.0, I48.1 |
| Congestive Heart Failure | 428 | I500, I501, I509 |
| Chronic Obstructive Pulmonary Disease | 491, 492, 496 | J41, J43, J44 |
| Coronary syndrome (excluding AMI) | 411-414 | I20, I22-I25 |
| Dementia | 290, 331 (OHIP) / 046.1, 290.0, 290.1, 290.2, 290.3, 290.4, 294, 331.0, 331.1, 331.5, F331.82 (DAD) | F00, F01, F02, F03, G30 |
| Diabetes | 250 | E08 - E13 |
| Hypertension | 401, 402, 403, 404, 405 | I10, I11, I12, I13, I15 |
| (Other) Mental Illnesses | 291, 292, 295, 297, 298, 299, 301, 302, 303, 304, 305, 306, 307, 313, 314, 315, 319 | F04, F050, F058, F059, F060, F061, F062, F063, F064, F07, F08, F10, F11, F12, F13, F14, F15, F16, F17, F18, F19, F20, F21, F22, F23, F24, F25, F26, F27, F28, F29, F340, F35, F36, F37, F430, F439, F453, F454, F458, F46, F47, F49, F50, F51, F52, F531, F538, F539, F54, F55, F56, F57, F58, F59, F60, F61, F62, F63, F64, F65, F66, F67, F681, F688, F69, F70, F71, F72, F73, F74, F75, F76, F77, F78, F79, F80, F81, F82, F83, F84, F85, F86, F87, F88, F89, F90, F91, F92, F931, F932, F933, F938, F939, F94, F95, F96, F97, F98 |
| Mood, anxiety, depression and other nonpsychotic disorders | 296, 300, 309, 311 | F30, F31, F32, F33, F34 (excl. F34.0), F38, F39, F40, F41, F42, F43.1, F43.2, F43.8, F44, F45.0, F45.1, F45.2, F48, F53.0, F68.0, F93.0, F99 |
| Osteoporosis | 733 | M81, M82 |
| Renal failure | 403, 404, 584, 585, 586, v451 | N17, N18, N19, T82.4, Z49.2, Z99.2 |
| **Stroke (excluding transient ischemic attack)** | 430, 431, 432, 434, 436 | I60-I64 |

**Table A3.**
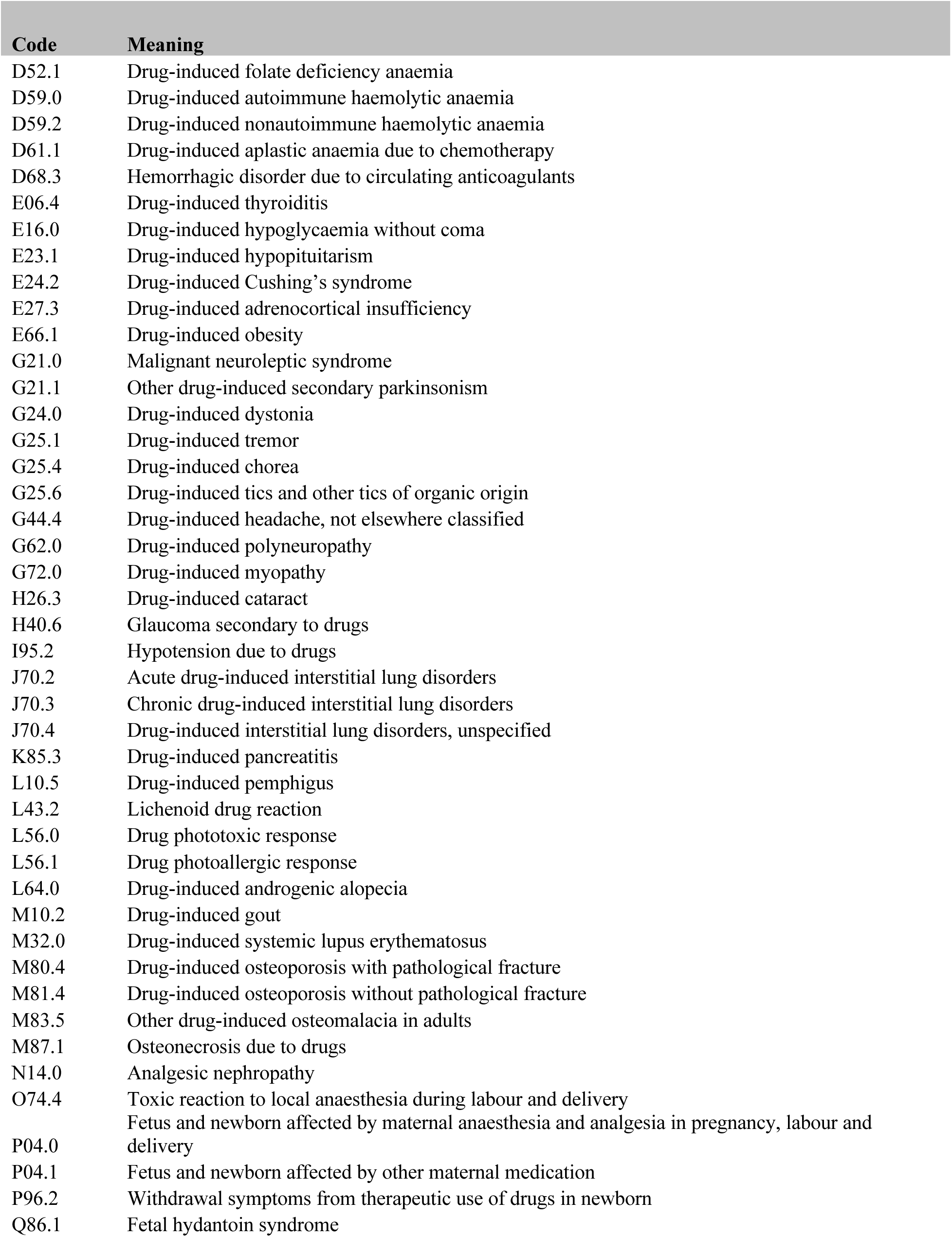
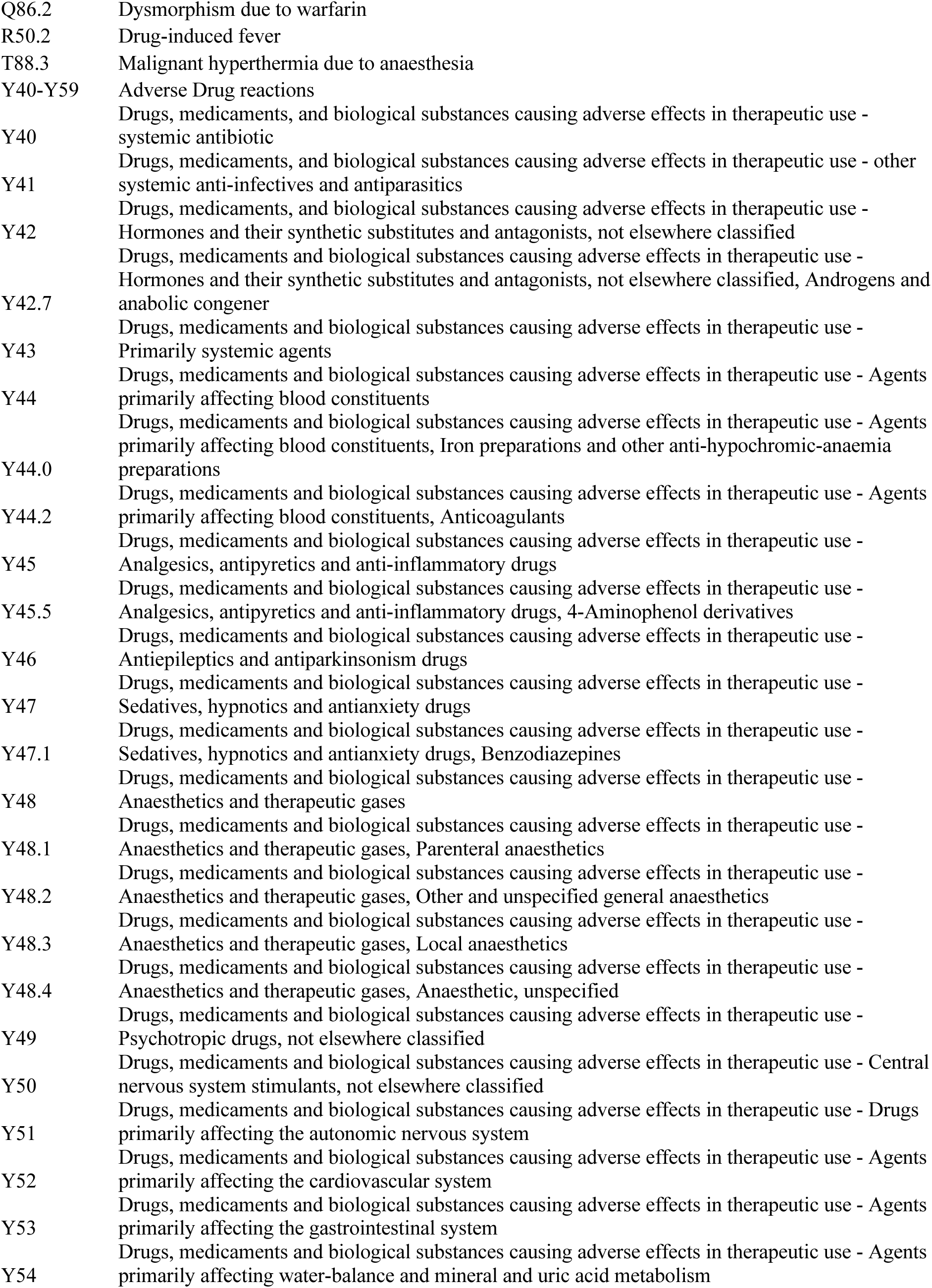
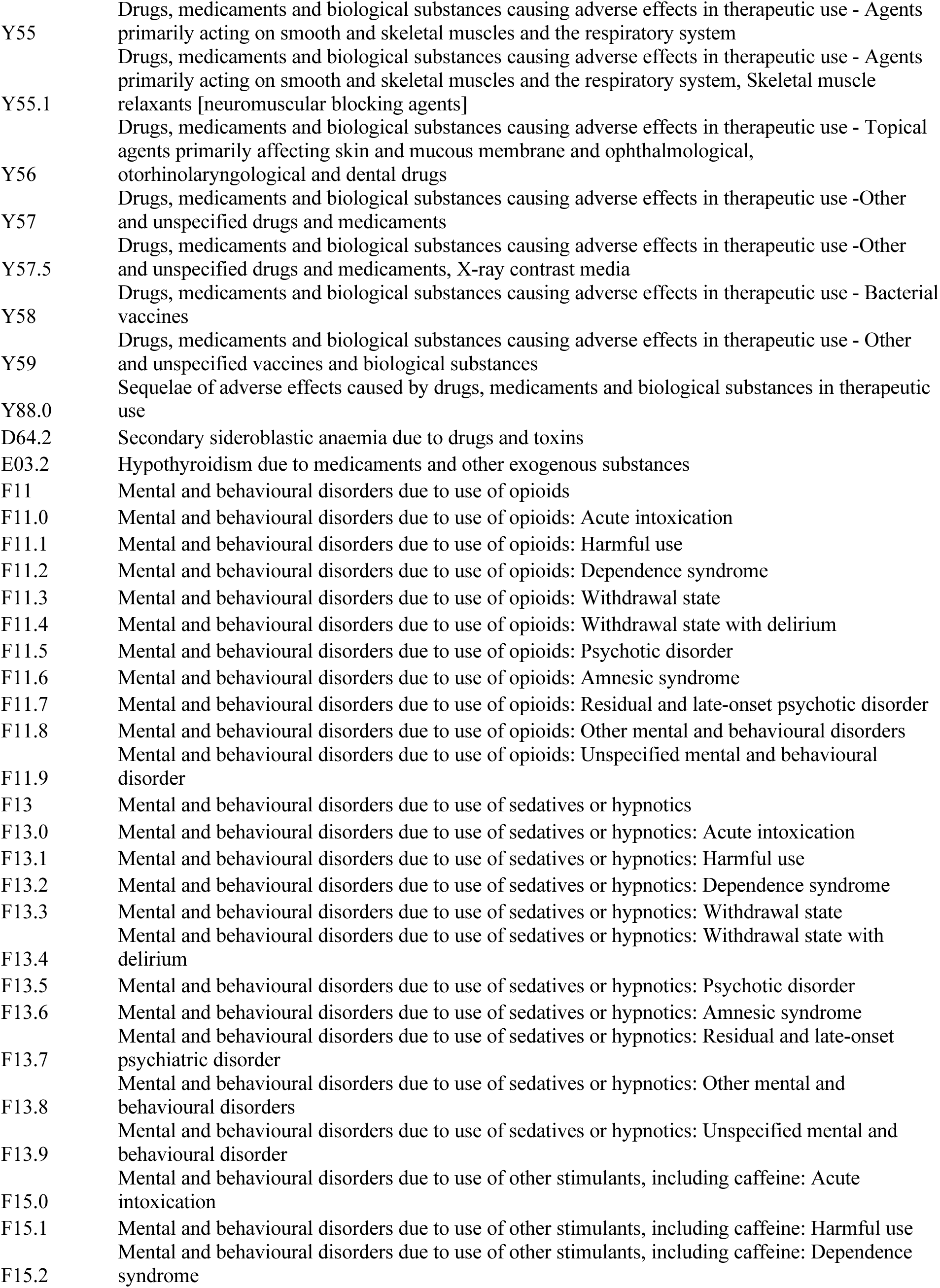
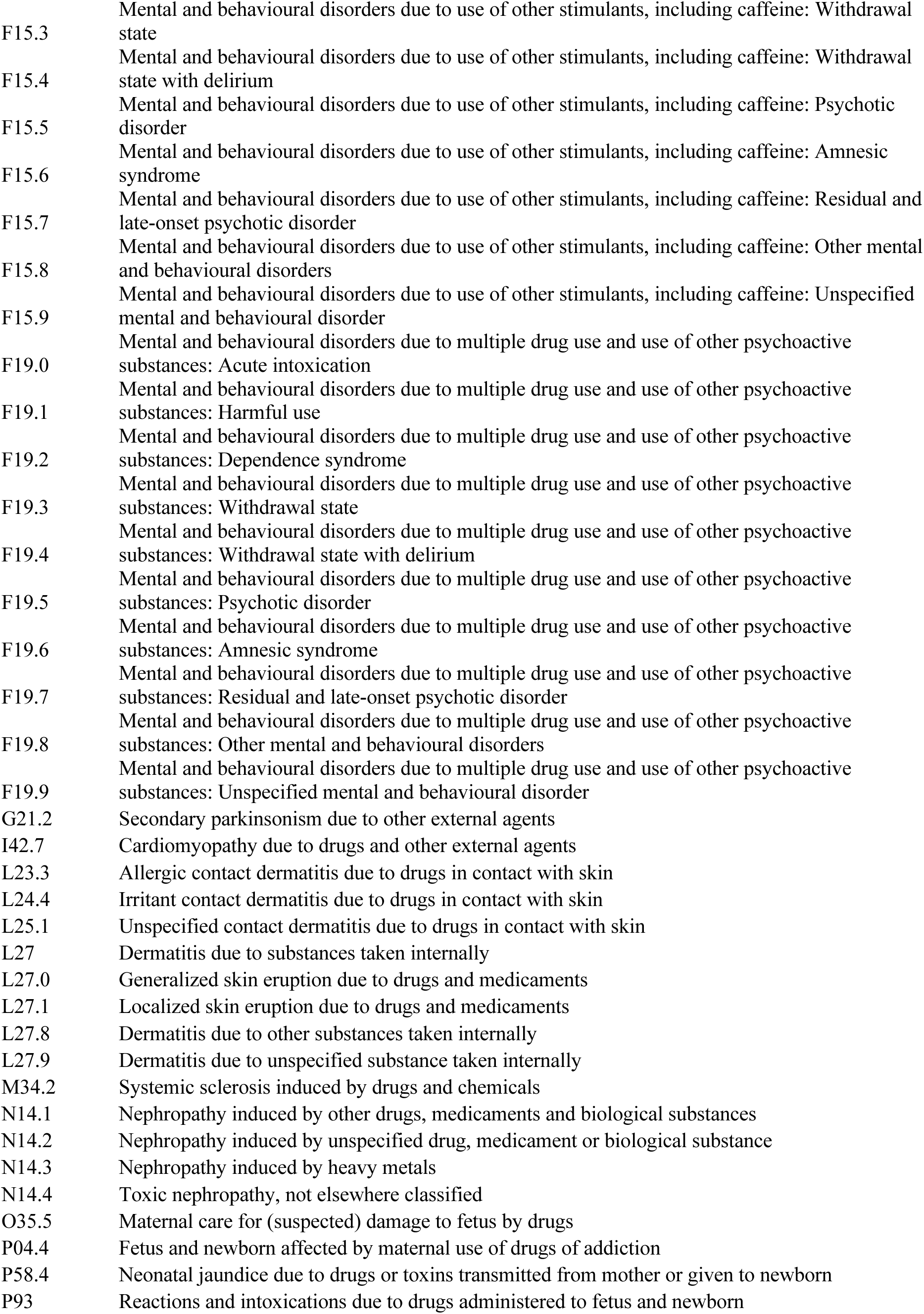
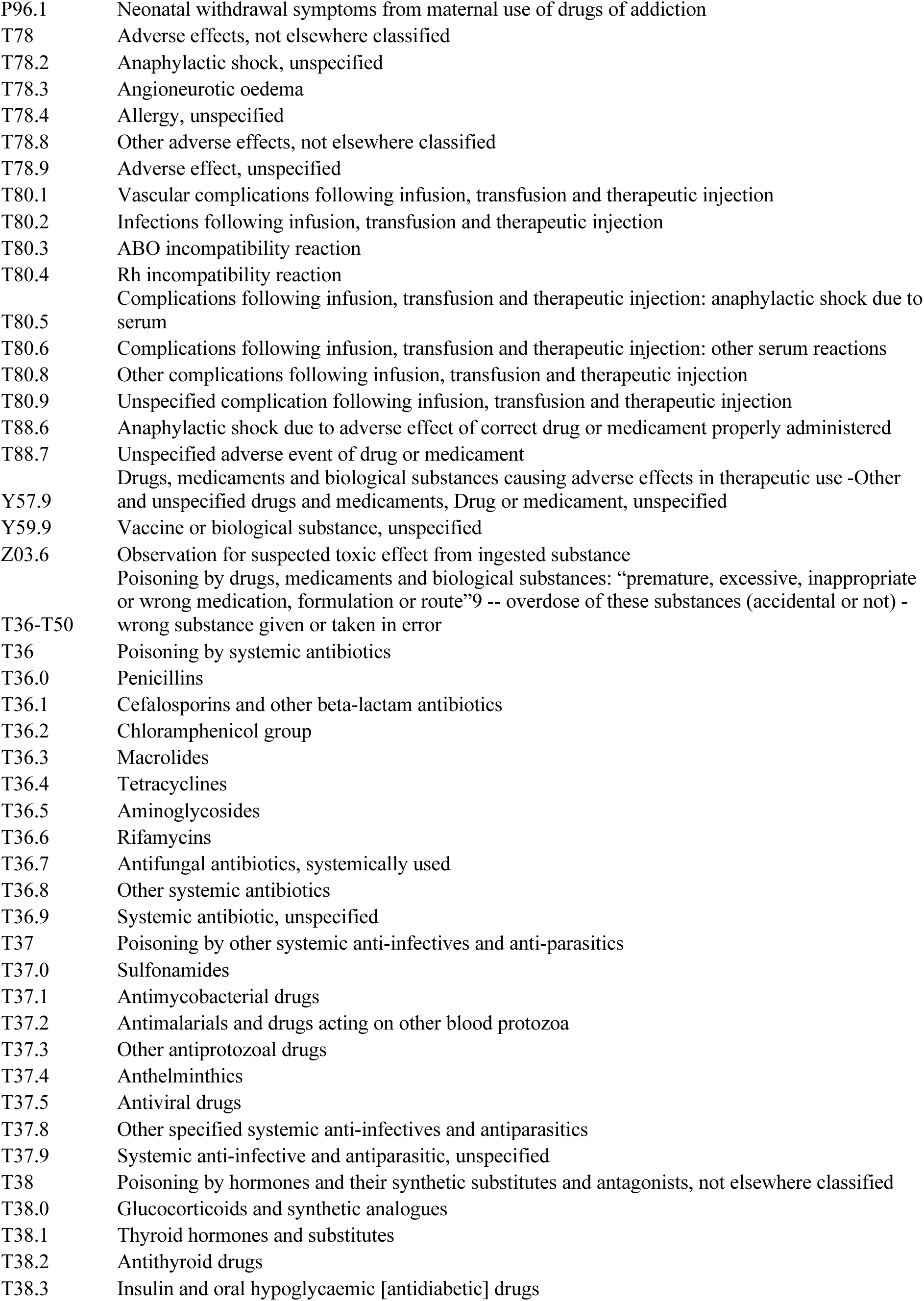
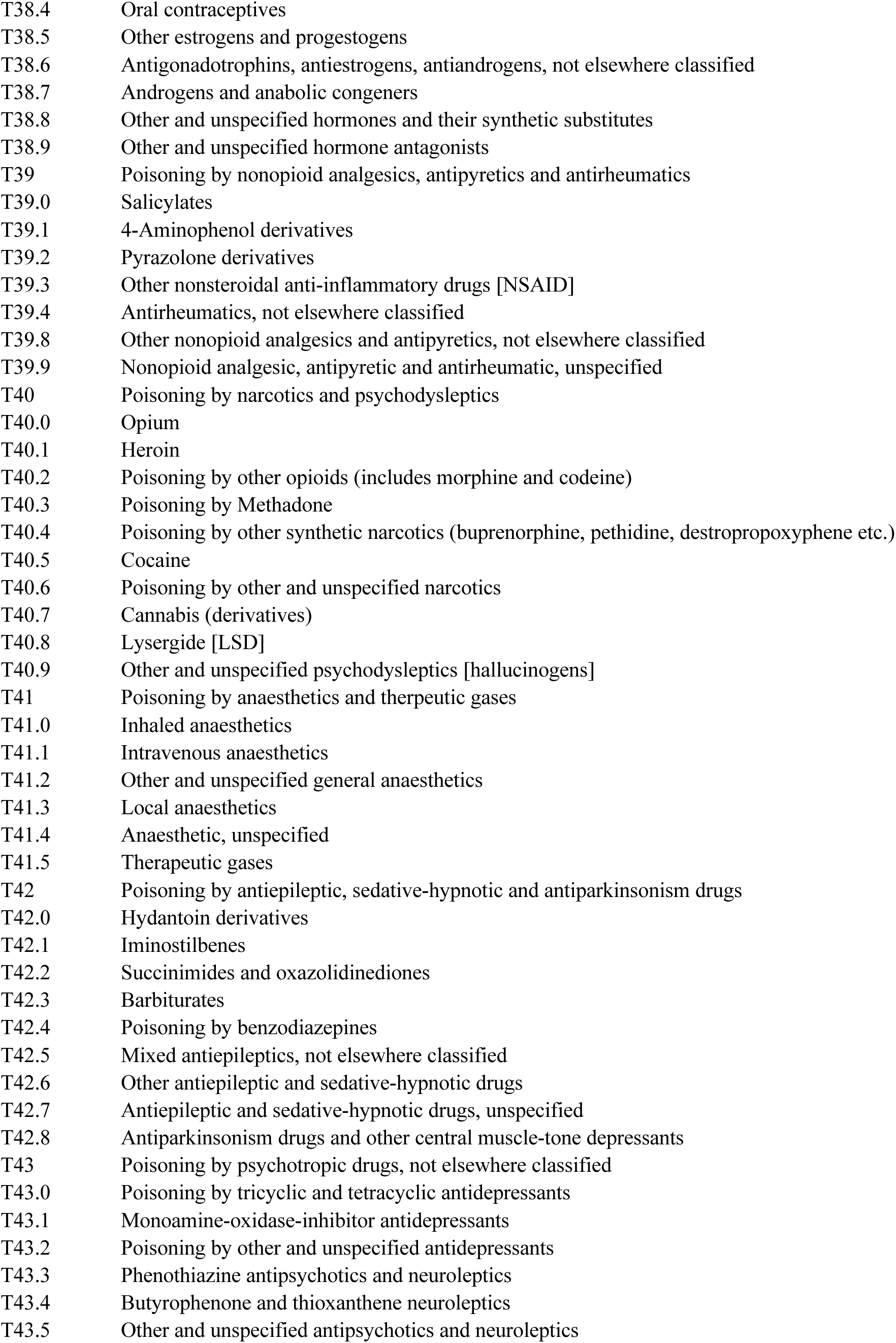
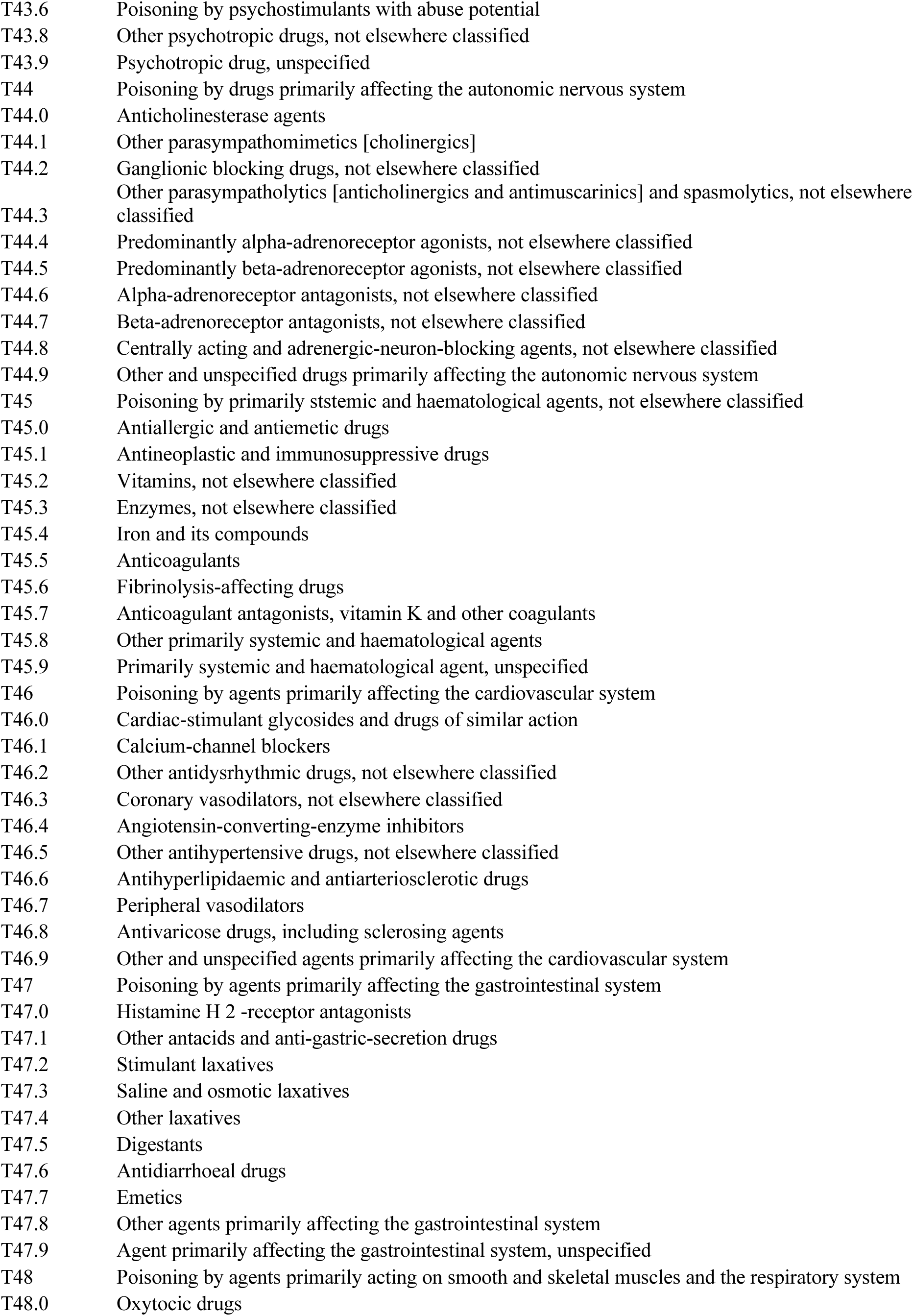
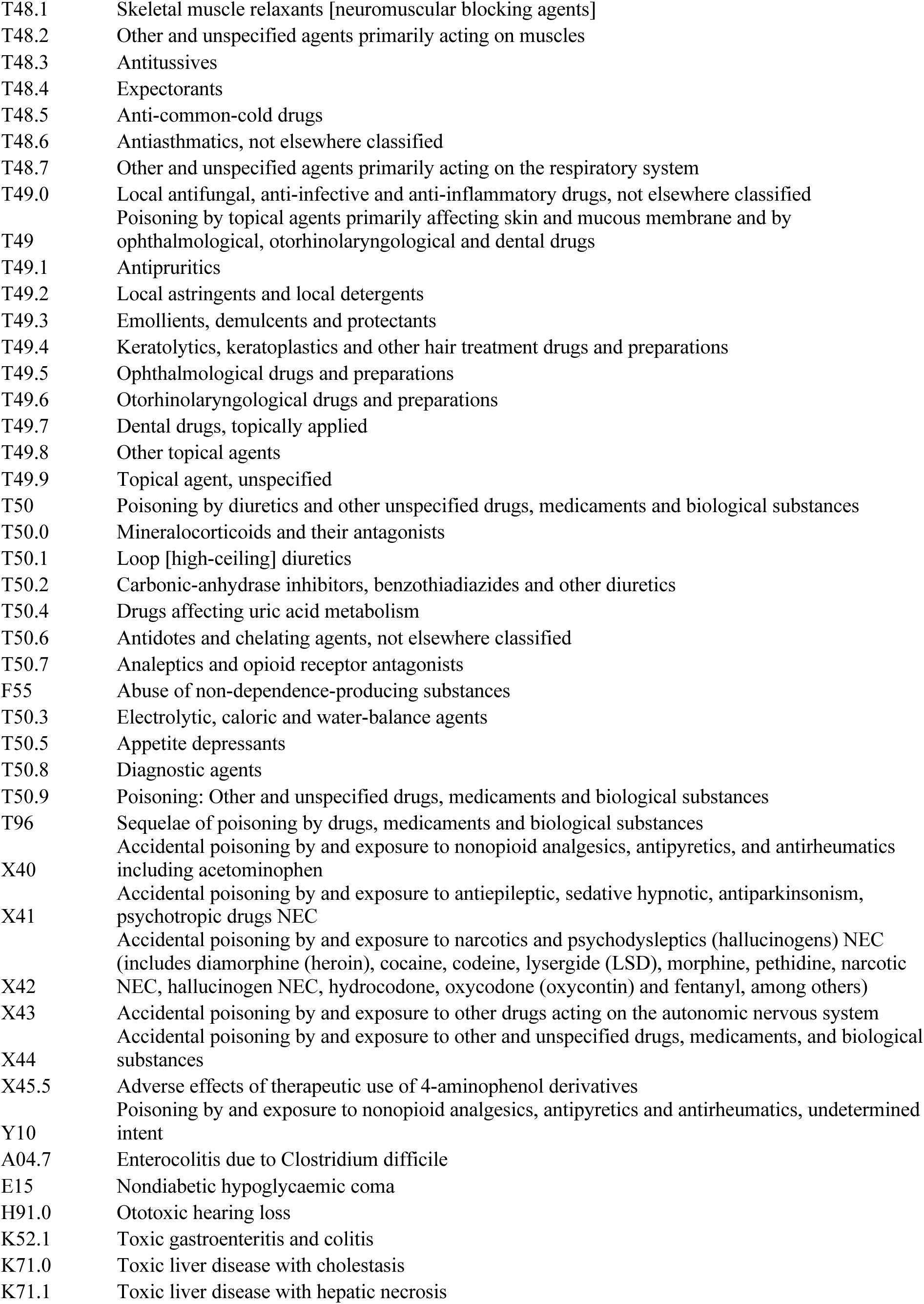
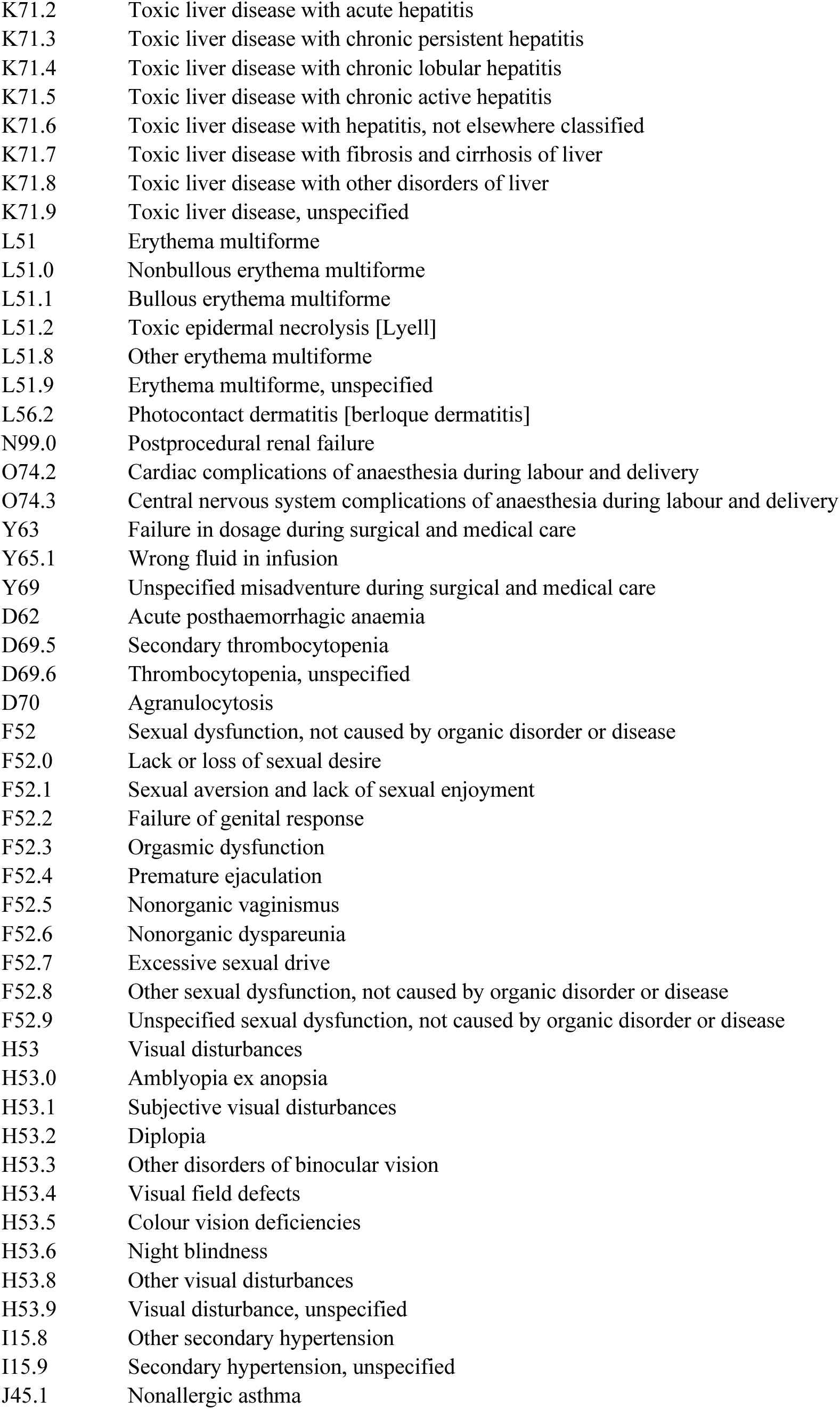
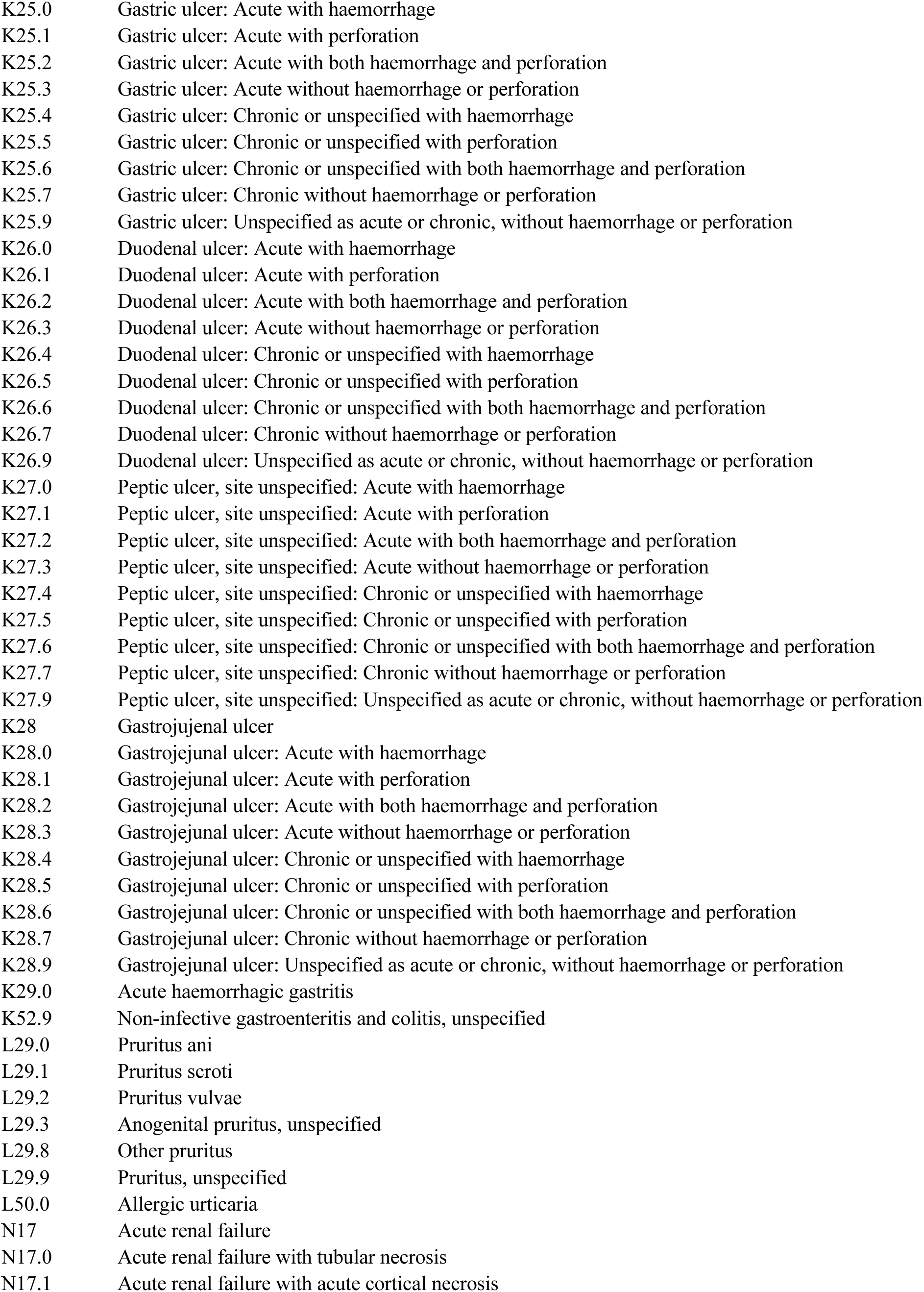
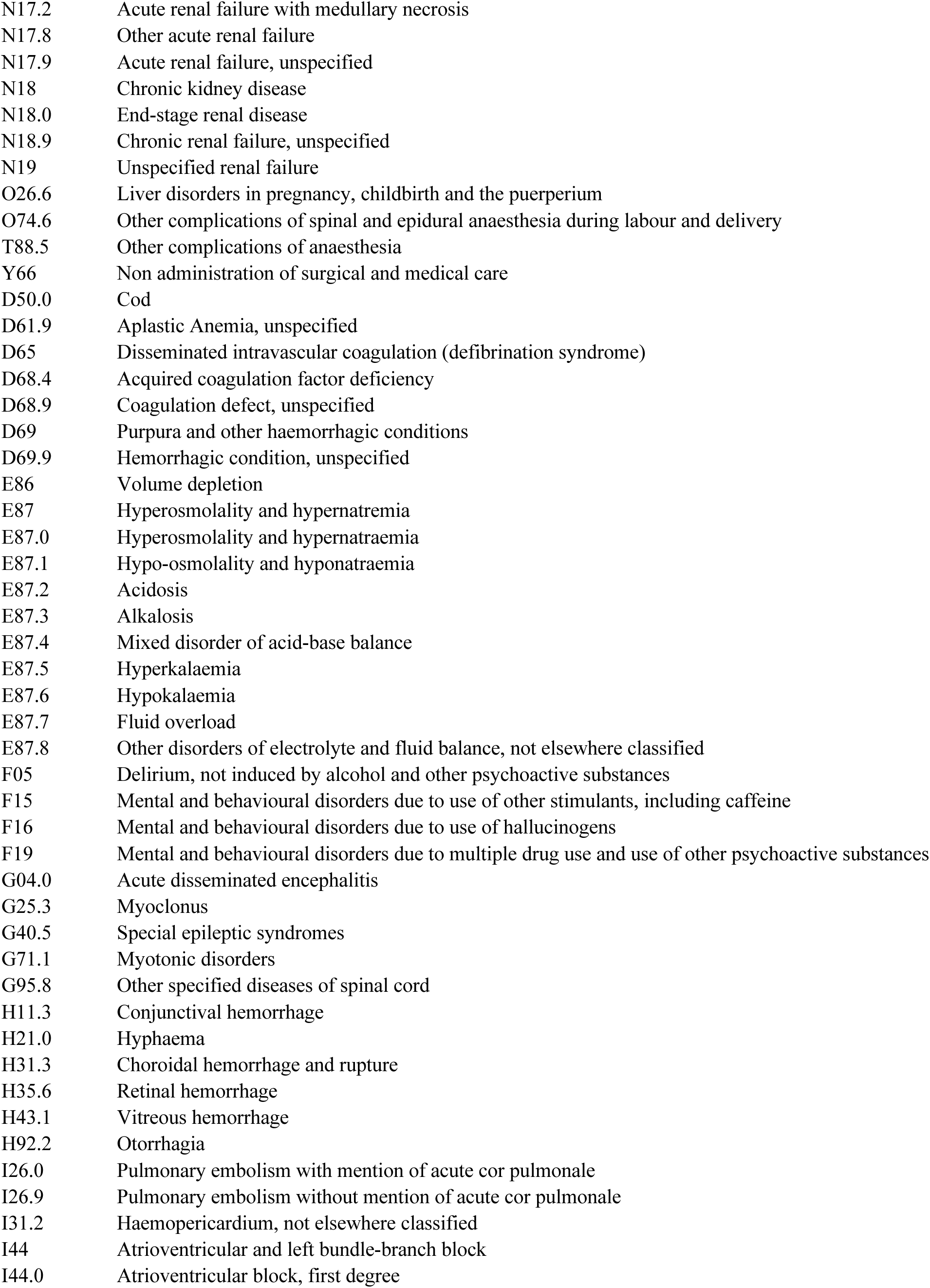
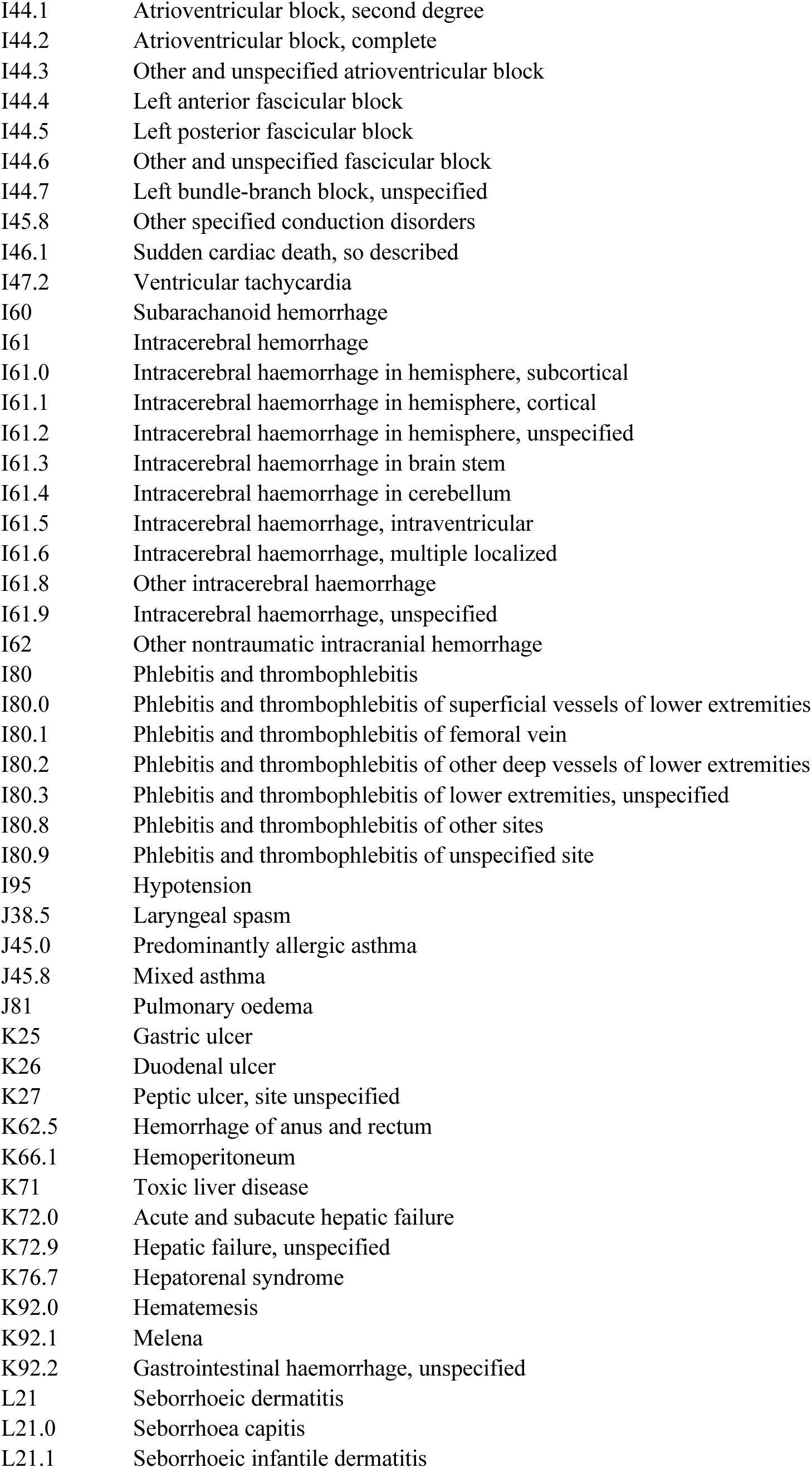
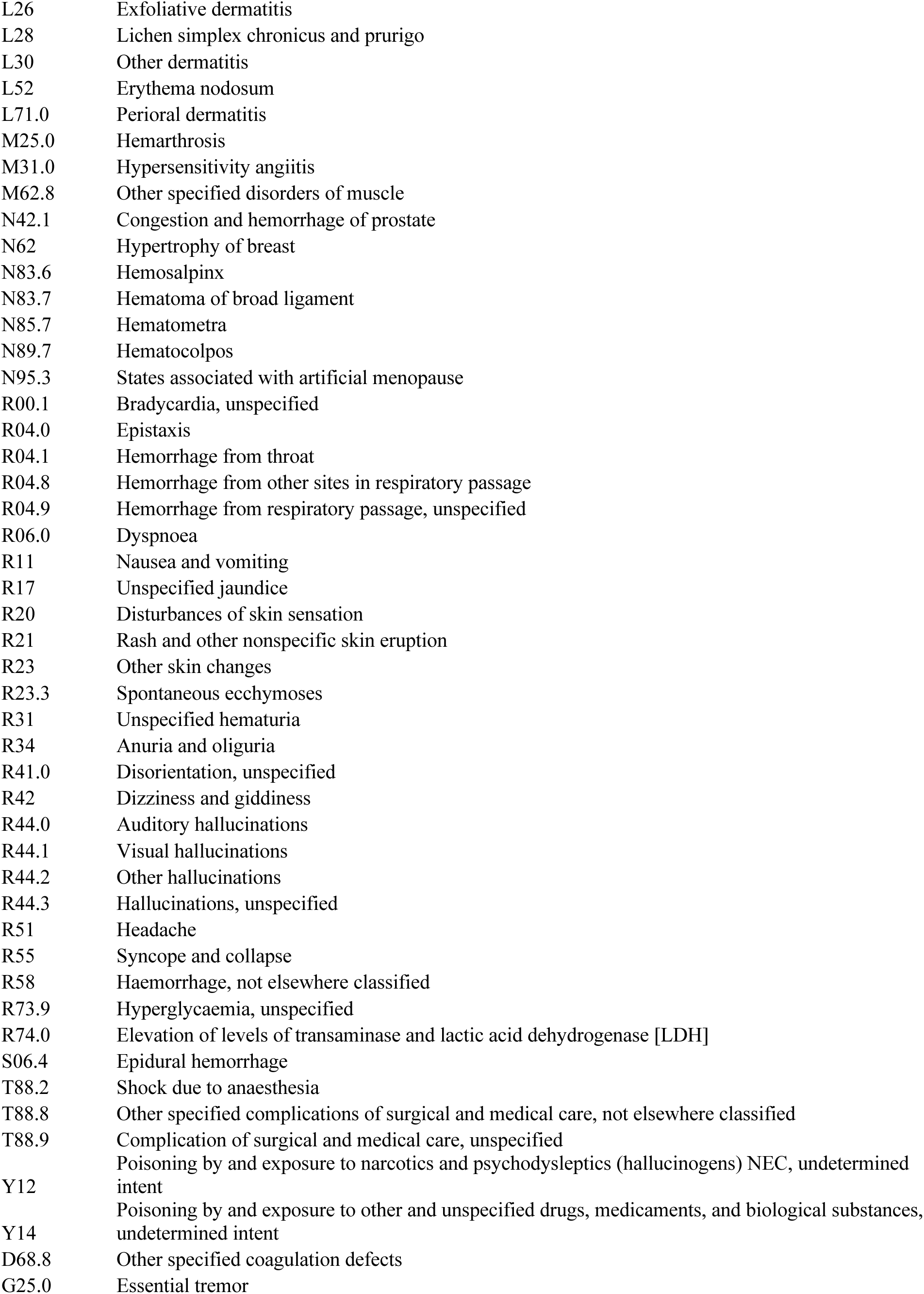
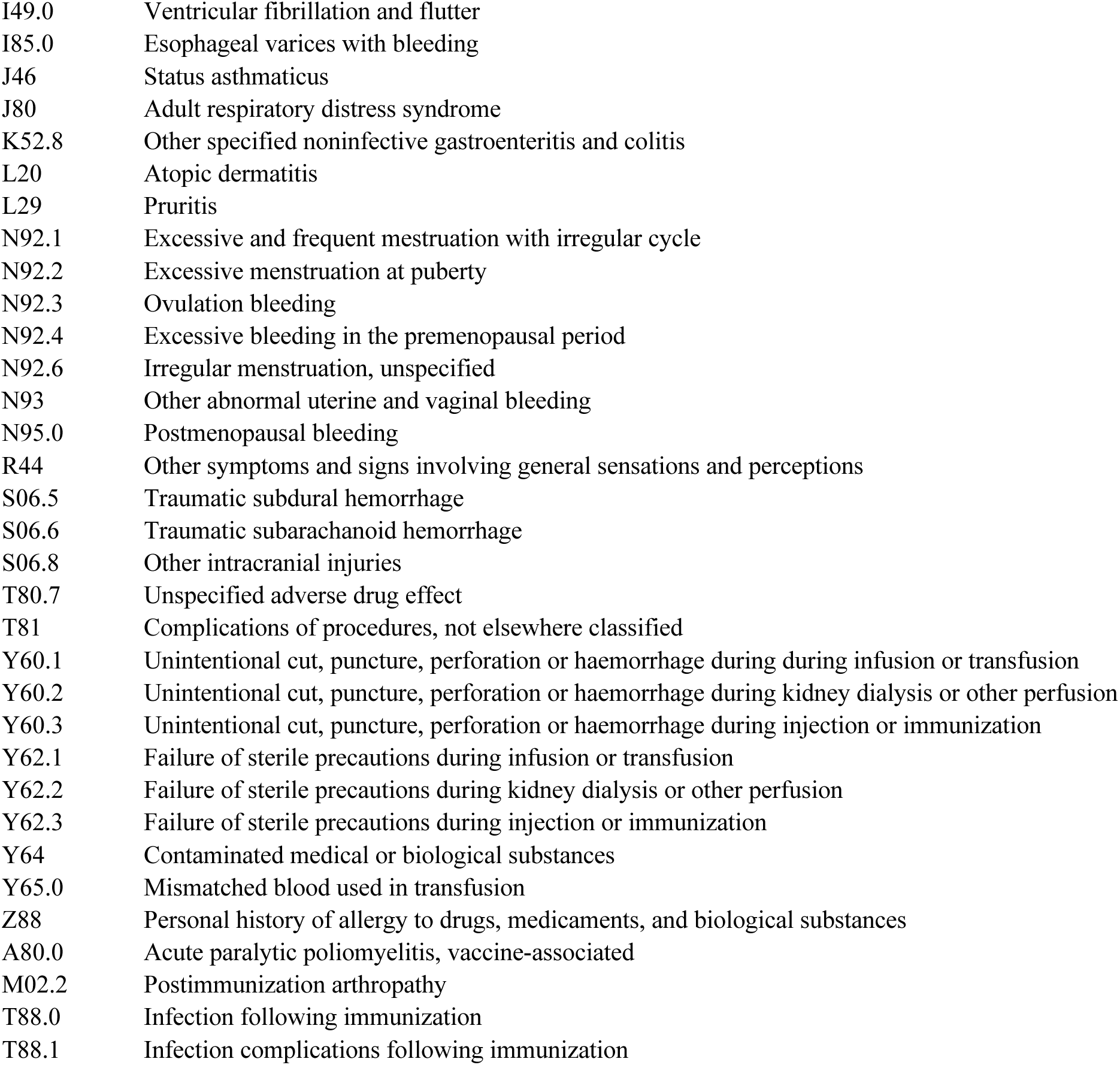
ICD 10 codes used to define adverse drug events (ADEs)

**Table A4.**
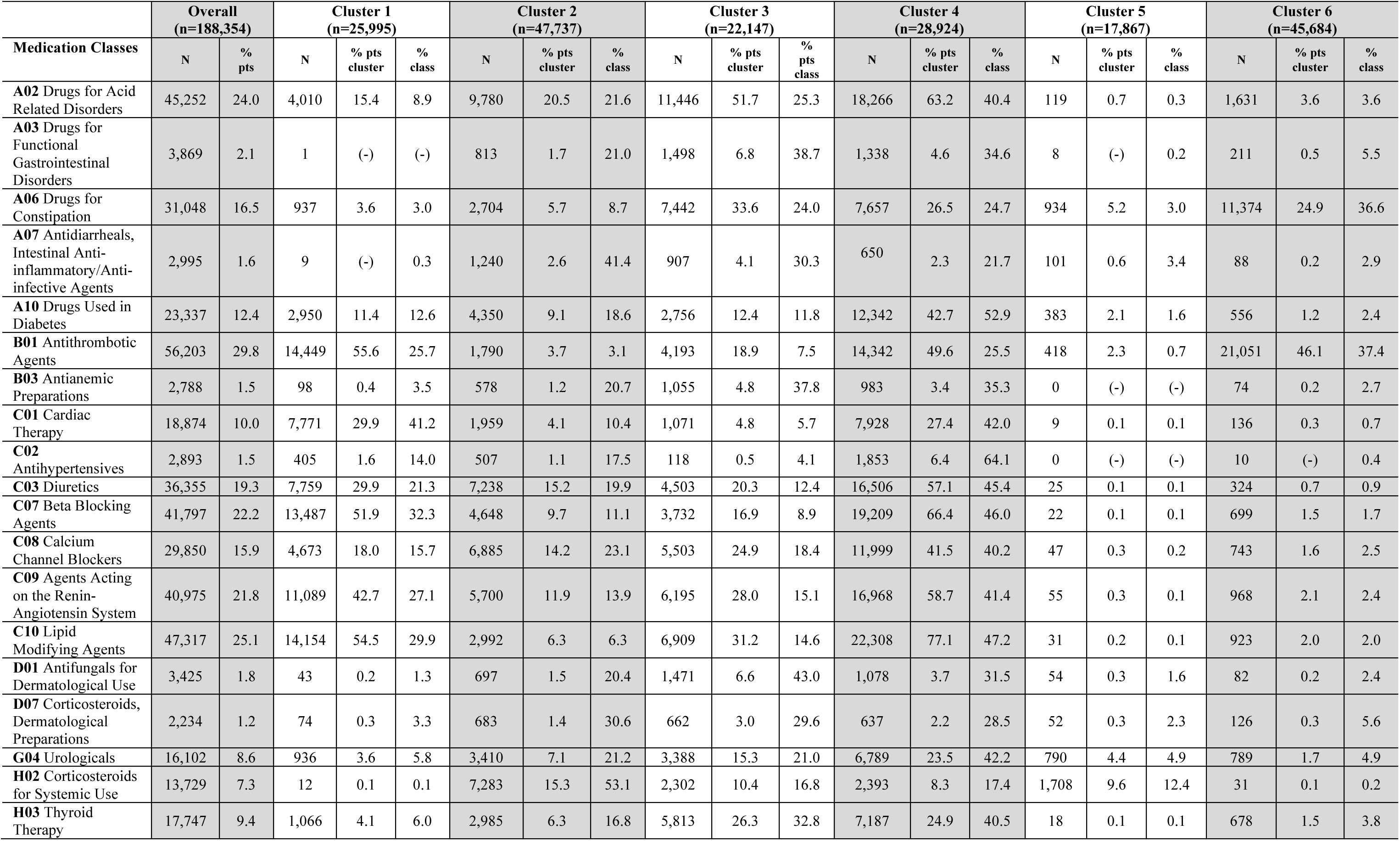
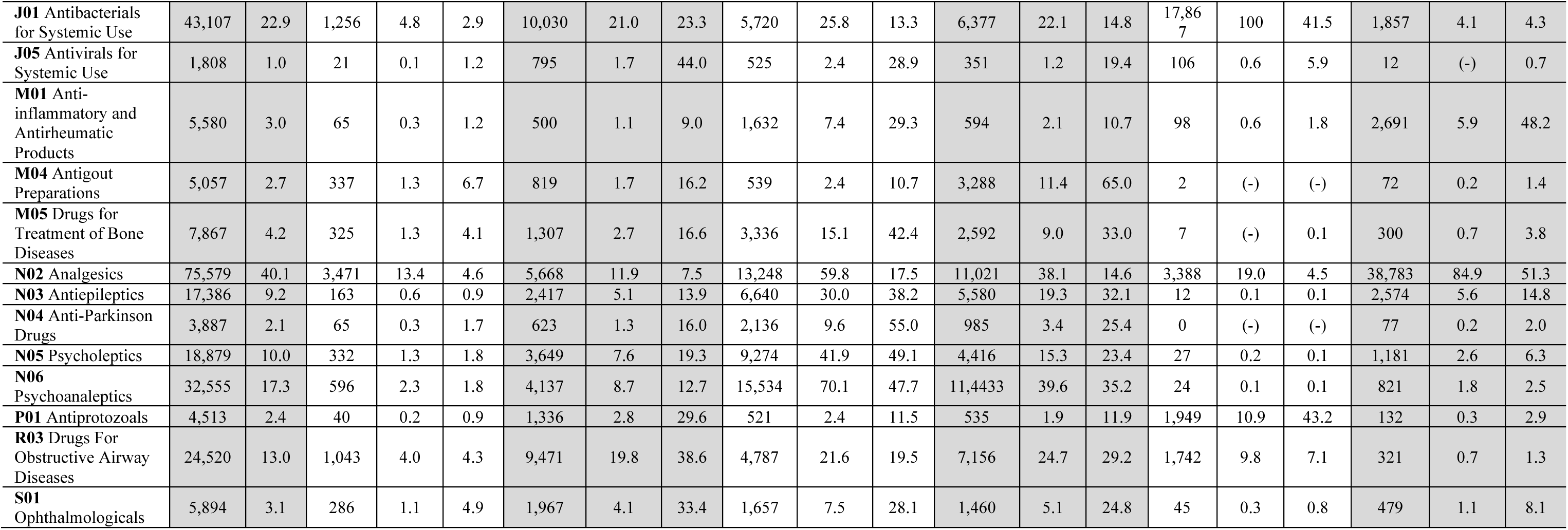
Medication Class Distributions Overall and According to Cluster

**Figure A1.**
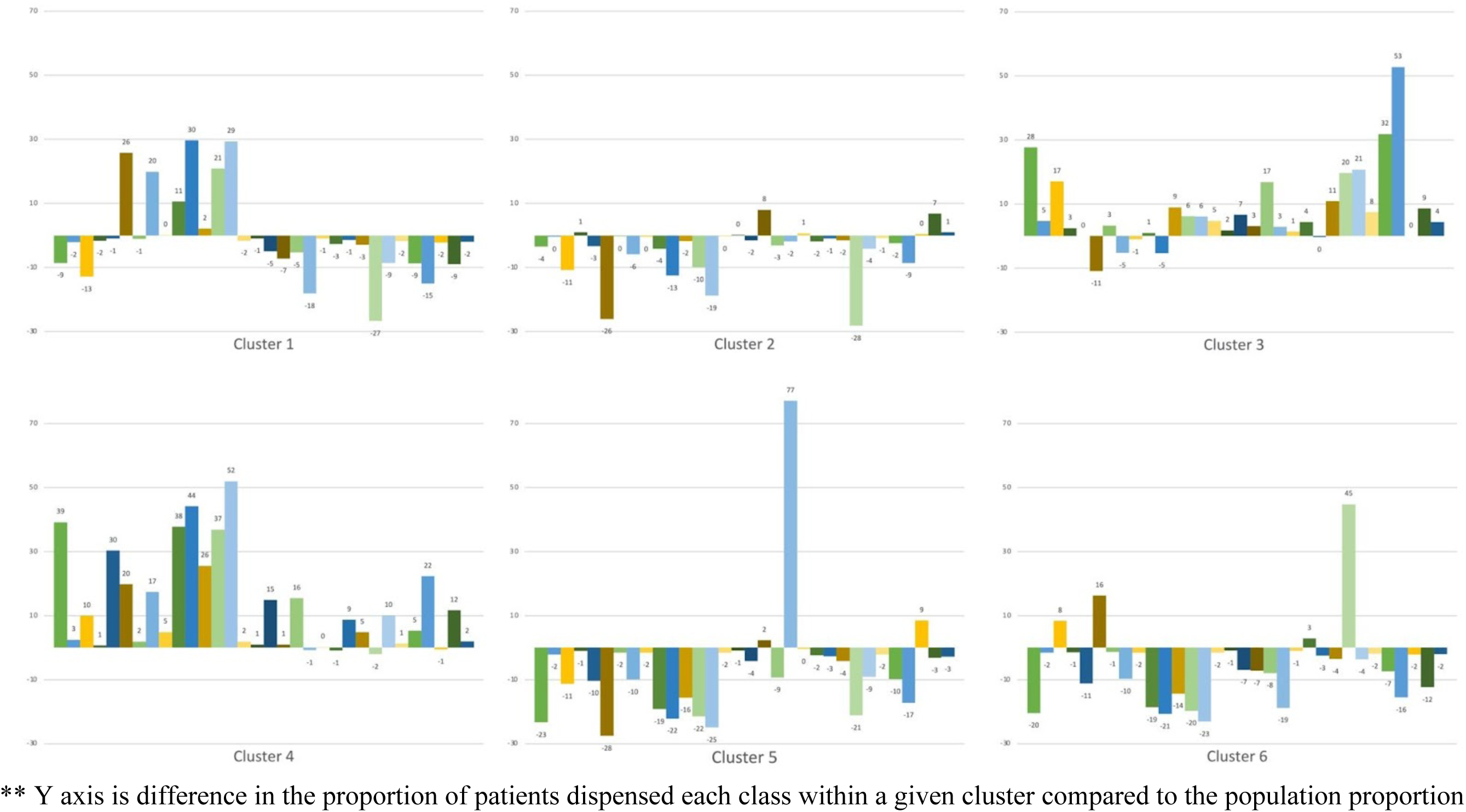
Difference in Prevalence of Medication Classes Dispensed in Each Cluster Compared to Study Population Prevalence

**Figure A2.**
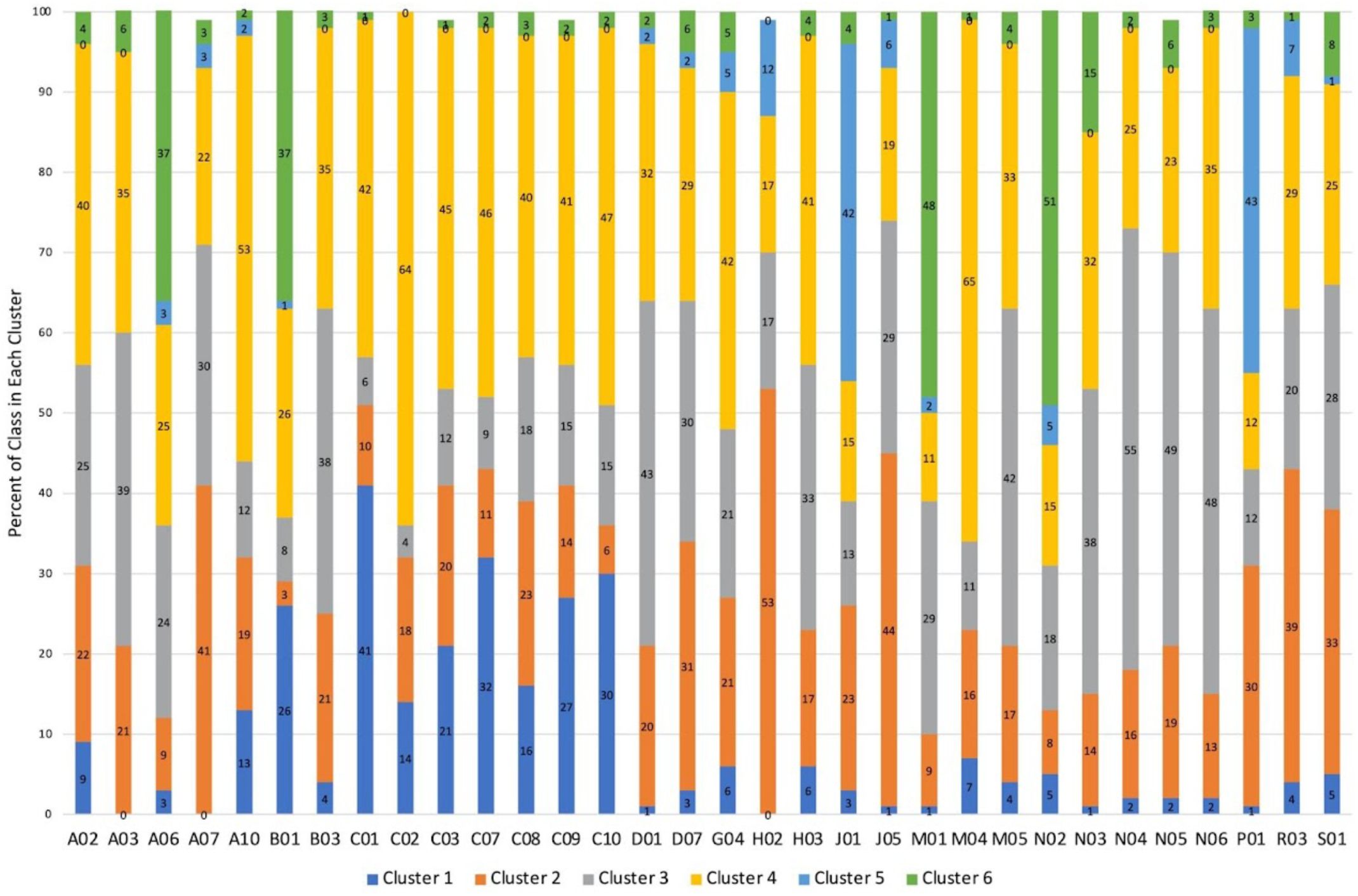
Prevalence of Cluster Membership within each Medication Class

**Table A5.** Sensitivity analysis-Association Between Medication Clusters and Risk of ED visits and mortality in 30-days Following Discharge Medication Cluster Crude

| Medication Cluster | Crude risk (%) | Adjusted OR* (95% CI) |
| --- | --- | --- |
| Cardiovascular | 24.2 | 0.89 (0.86-0.92) |
| Respiratory | 29.5 | 1.07 (1.05-1.10) |
| Complex care needs | 25.3 | 0.80 (0.77-0.83) |
| Cardiovascular and metabolic | 27.1 | 0.80 (0.77-0.82) |
| Infection | 33.2 | 1.33 (1.28-1.38) |
| Surgical | 24.0 | 1.21 (1.17-1.25) |

\* Reference is all other clusters (e.g., cardiovascular vs not)

**Table A6.**
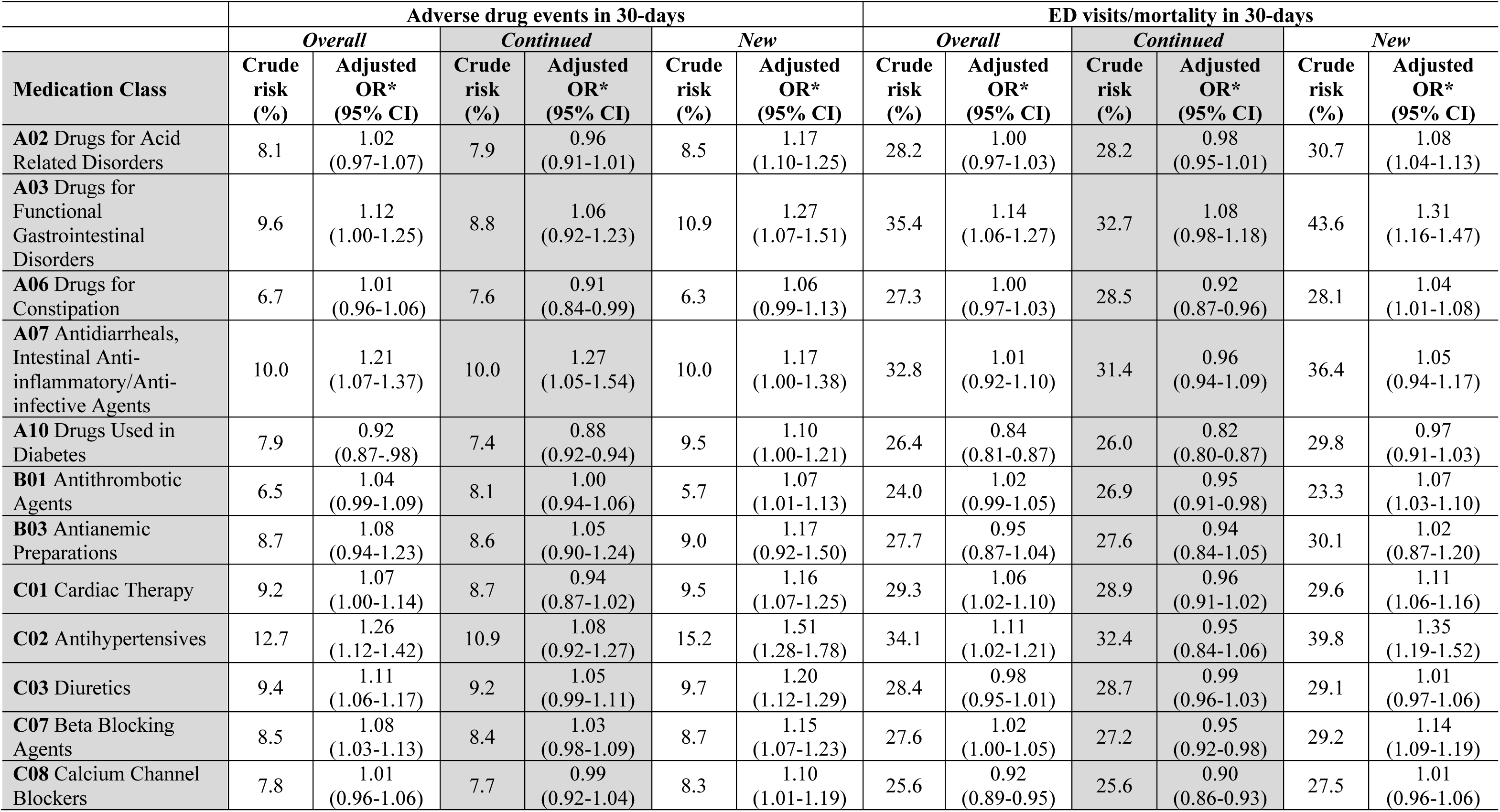
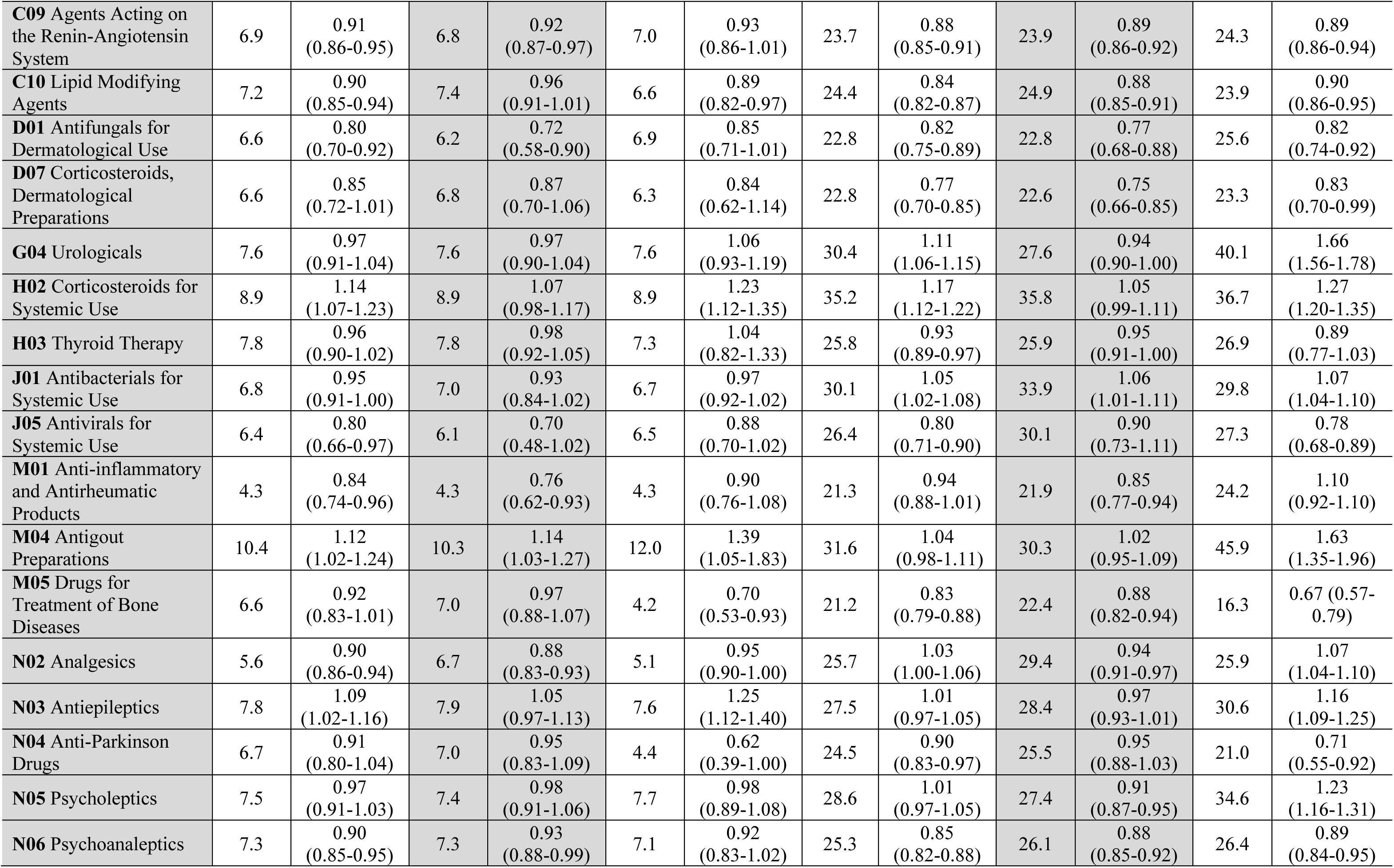
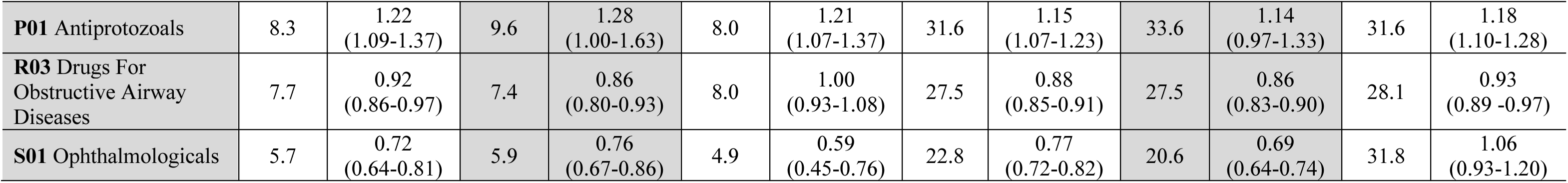
Association between medication class and ED visits/mortality and ADEs in 30-days following discharge

